# T Cell Receptor repertoire analysis reveals antigenic convergence and immunotherapeutic opportunities in Prostate Cancer

**DOI:** 10.64898/2026.06.12.26355376

**Authors:** Abatino Antonio, Giordano Caterina, Isdraele Romano Lisa, Fiume Giuseppe, Sarubbi Maria Chiara, Malanga Donatella, Amodeo Antonio, Alba Stefano, Alberto Piana, Cristian Fiori, Damiano Rocco, Cantiello Francesco, Crocerossa Fabio, Cuda Giovanni, Gallo Raffaella, Aversa Ilenia, Palmieri Camillo

## Abstract

**Background:** The T-cell receptor β (TCRβ) repertoire reflects antigen-driven adaptive immune responses and provides insight into tumor–immune interaction. In prostate cancer (PCa), the immunosuppressive tumor microenvironment limits effective T-cell activation, and the antigenic drivers shaping intratumoral TCR repertoires remains poorly defined. This study aimed to characterize matched tumor and peripheral TCRβ repertoires from treatment-naïve PCa patients and to identify shared clonotypes and antigenic specificities associated with disease severity.

**Methods:** Next-generation sequencing was used to profile TCRβ repertoires from matched tumor biopsies and peripheral blood mononuclear cells (PBMCs) obtained from treatment-naïve PCa patients. Repertoires clonality, diversity, and was assessed using established metrics. Antigenic convergence was evaluated using GLIPH2 to identify shared CDR3β motifs and predicted tumor-associated antigen (TAA) recognition. Preliminary functional validation was performed using IFN-γ ELISpot, T-cell expansion, and single-cell RNA sequencing in an independent cohort of PCa patients.

**Results:** Tumor-derived TCRβ repertoires displayed reduced richness and increased clonality compared with PBCMs, consistent with local antigen-driven expansion. High-grade tumors demonstrated greater interpatient clonotype sharing and motif-level convergence, indicative of recognition of common TAAs. GLIPH2 analysis associated expanded clonotypes with epitopes derived from prostate-specific G-protein coupled receptor (PSGR), prostate-specific membrane antigen (PSMA), and prostate-specific antigen (PSA). Functional validation confirmed that peptide pools containing PSGR-, PSMA-, and Transglutaminase-4 (TGM4)-derived epitopes induced IFN-γ production and antigen-specific T-cell proliferation in vitro. Single-cell RNA sequencing demonstrated an enhanced activation and cytotoxic transcriptional profile in CD3⁺ T cells following stimulation with the peptide pool.

**Conclusions:** These findings reveal an oligoclonal, antigen-driven intratumoral TCRβ landscape and identify PSGR, PSMA, and TGM4 as immunogenic, potentially actionable targets. Integration of TCR profiling with antigen discovery pipelines may support the development of TCR-based biomarkers and precision immunotherapeutic strategies in prostate cancer.

## BACKGROUND

Prostate cancer (PCa) is a major global health issue, with an estimated 1.47 million new cases and 375,000 deaths worldwide in 2022 and remains the most diagnosed malignancy and the second leading cause of cancer-related death among men [1]. Treatment depends on risk stratification, prostate-specific antigen (PSA) levels, and life expectancy. For localized disease, surgery or radiotherapy represent the main treatment options, whereas androgen deprivation therapy (ADT), often combined with additional systemic treatments, remains a cornerstone of therapy for patients with advanced or metastatic disease [2], [3]. Despite initial treatment responses, disease progression and the development of castration resistance remain major clinical challenges. Metastatic castration-resistant prostate cancer (mCRPC) is an incurable condition associated with substantial mortality and limited long-term therapeutic options. Consequently, considerable efforts have been directed toward identifying novel molecular targets and therapeutic strategies beyond androgen receptor signaling [4]. Immunotherapy has achieved limited success in PCa; Sipuleucel-T, an autologous dendritic cell-based vaccine, designed to elicit an immune response against prostatic acid phosphatase (PAP), remains the only FDA-approved cellular immunotherapy for PCa [5], [6]. However, its clinical benefit is limited to a modest improvement in overall survival without a significant impact on disease progression [7]–[9]. Similarly, other vaccine-based strategies [10], [11] and immune checkpoint inhibitors targeting CTLA-4 and PD-1/PD-L1 [12]–[25] have shown limited activity in unselected patients with mCRPC. Nevertheless, clinical benefit has been observed in biomarker-defined subsets, including tumors with T-effector-high or PD-L1-positive immune profiles and those characterized by deficient mismatch repair or high microsatellite instability (dMMR/MSI-H) [12], [16], [21], [26], [27].The overall low responsiveness of PCa to current immunotherapies is largely attributed to its classification as an “immunologically cold” tumor, characterized by a profoundly immunosuppressive tumor microenvironment, limited T-cell infiltration, and generally low tumor mutational burden, which together contribute to poor immunogenicity [21], [28]–[30] . To overcome these limitations, increasing attention has been directed toward alternative T-cell-based therapeutic strategies. CAR-T cell therapies targeting prostate-specific membrane antigen (PSMA) and prostate stem cell antigen (PSCA) are under investigation and have shown preliminary evidence of safety and antitumor activity [28], [31]. In parallel, T-cell engagers and bispecific antibodies targeting prostate cancer-associated antigens, including PSMA, Six-Transmembrane Epithelial Antigen of the Prostate 1 (STEAP1), and kallikrein-related peptidase 2 (KLK2), have entered early-phase clinical development and demonstrated initial signs of clinical activity [32], [33]. These developments underscore the growing interest in harnessing T-cell-mediated immunity in PCa and highlight the need to better characterize endogenous antitumor T-cell responses and their antigenic targets.

In this context, analysis of the T-cell receptor (TCR) repertoire provides a powerful approach to investigate adaptive immune responses in cancer. Each T cell expresses a unique TCR sequence that defines its clonotype, a population of T cells sharing the same antigen specificity. The collection of all clonotypes within a biological compartment, such as peripheral blood or tumor tissue, constitutes the TCR repertoire and provides a molecular fingerprint of immune history and activity. In solid tumors, repertoire composition, clonal expansion and clonotype dynamics can reflect antigen-driven selection and T-cell responses to tumor-associated antigen or neoantigens [34], [35]. Advances in next-generation sequencing (NGS) have enabled high-resolution profiling of TCR repertoires, supporting their investigation as biomarkers of antitumor immune responses, treatment response, and clinical outcome. Moreover, integration of repertoire sequencing with computational approaches may enable the identification of candidate antigen-associated TCR groups and the prioritization of clonotypes for subsequent functional validation. Diversity and clonality metrics derived from repertoire sequencing have shown diagnostic, prognostic, and predictive relevance across multiple cancer types [34], [36], [37].

In PCa, despite its immunologically cold microenvironment, relatively few studies have investigated matched intratumoral and peripheral TCR repertoires. Available evidence has revealed expanded intratumoral T-cell clonotypes, supporting the presence of antigen-driven T-cell responses and potentially tumor-reactive T-cell populations [37], [38]. However, the relationship between intratumoral and circulating repertoires and the antigenic targets associated with shared or expanded clonotypes remain incompletely characterized.

In this study, we characterized matched intratumoral and peripheral TCRβ repertoires from ten treatment-naïve PCa patients. The objectives were twofold: (i) to quantify the overlap between peripheral and intratumoral repertoires, thereby investigating whether peripheral blood may capture tumor-associated immune signatures and represent a potential source of candidate tumor-reactive T cells; and (ii) to identify candidate antigen-associated specificity groups among shared and intratumoral clonotypes through computational analyses and *in vitro* peptide-stimulation assays, with the aim of prioritizing putative tumor-associated antigens and TCR candidates for subsequent functional validation.

## MATERIALS AND METHODS

### Study design and characteristics of the enrolled patients

Tumor biopsies and peripheral venous blood were collected from 26 prostate cancer (PCa) patients at Romolo Hospital Clinic (Crotone, Italy) as part of a collaboration with the Magna Graecia University of Catanzaro (Catanzaro, Italy). Patients (PSA 12.15 ± 7.34 ng/mL) were eligible for inclusion if they were male and undergoing prostate biopsy due to a clinical suspicion of PCa. This suspicion was based on PSA levels, abnormal digital rectal examination (DRE), and/or a family history of PCa. All patients had to be suitable for the full biopsy protocol, including transrectal ultrasound. Exclusion criteria included a history of other malignancies, evidence of urinary tract infections, or regular use of immunosuppressive drugs. All patients provided written informed consent. Histological classification included acinar adenocarcinoma, ductal adenocarcinoma, and high-grade prostatic intraepithelial neoplasia (PIN).

All materials derived from diagnostic surplus samples, namely, tissue fragments remaining after routine histopathological analyses and residual blood collected for pre-biopsy laboratory testing. Of note, all patients were not under hormonal treatment for Benign Prostatic Hyperplasia. For the in vitro functional validation assays, peripheral venous blood samples were obtained from a second cohort composed of 18 patients with a history of PCa, monitored through PSA levels (mean 17.38 ± 11.82, range: 4.26-20.8 ng/mL) and 4 healthy donors.

### Peripheral blood mononuclear cells purification

Peripheral venous blood was collected in EDTA vacutainer tubes, and peripheral blood mononuclear cells (PBMCs) were isolated by density gradient using Histopaque-1077 gradient centrifugation (code 10771; Sigma-Aldrich, Darmstadt, Germany), according to the manufacturer’s instructions and as previously reported[39] . Isolated PBMCs were cryopreserved and stored in liquid nitrogen until use.

### RNA extraction and quality control

RNA from PBMC samples was isolated using the TRIzol reagent (Thermo Fisher Scientific, Milan, Italy) as described before [40] . Tumor tissues were stored in RNA later solution (AM702; ThermoFisher Scientific) at -80°C until use. Tumor biopsies (4–5 mm) were first homogenized in 0.5–1 mL of TRIzol Reagent and stored at −80 °C for 2–3 days to ensure complete tissue dissociation prior to extraction. After extraction, RNA was subjected to quality assessment by determining the RNA Integrity Number (RIN). RIN values were measured with the TapeStation 4200 system (Agilent Technologies) using ScreenTape RNA High Sensitivity (codes 5067 5579, 5067-5580, 5067-5581; Agilent Technologies) following the manufacturer’s instructions. Only RNA samples with a RIN ≥ 5.5 were selected, reverse transcribed through the InvitrogenTM SuperScript IV VILO Master Mix (ThermoFisher Scientific) according to the manufacturer’s instructions, and used for TCRβ library construction.

### TCR sequencing, raw data processing and single-cell analysis

TCRβ libraries were prepared for NGS sequencing using the Oncomine™ TCR Beta-LR Assay (ThermoFisher Scientific), as previously described [34]. RNA from PBMCs was reverse-transcribed using SuperScript IV VILO Master Mix (ThermoFisher Scientific), and amplified with AmpliSeq primers targeting the framework region 1 (FR1) and constant (C) regions of TCRβ, generating 330 bp amplicons. Library preparation was manually performed using the Ion AmpliSeq Kit for Chef DL8 (ThermoFisher Scientific), and cDNA concentration was measured with TapeStation 2200 system using the High Sensitivity DNA Assay (codes 5067-5584, 5067-5585; Agilent Technologies, Milano, Italy). Barcoded libraries were diluted to 25 pM and loaded onto the Ion Chef™ system for emulsion PCR, enrichment, and sequencing on the Ion S5 530 chip.

Post-sequencing, raw data were analyzed with the Ion Torrent Suite Software. Alignment of V, D and J segments, CDR3 identification, and assembly of reads into clonotypes were performed with MiXCR (v.4.1.2) using the built-in preset pipeline for the "Oncomine TCR Beta LR Assay" [41] . Downstream analyses were restricted to productive TCRβ rearrangements. Clonotypes were defined by unique CDR3 amino acid sequences, and repertoire analyses were performed using productive clonotypes without additional abundance filtering, unless otherwise specified. TCR repertoire analysis was performed using the Immunarch 1.0.0 R package [42] . MiXCR output files were imported into R using the repLoad function. Repertoire characteristics, including clonotype abundance and repertoire size, were explored using repExplore. Diversity metrics (Richness, normalized Richness, Shannon entropy, Pielou’s evenness, and Simpson index) were calculated using repDiversity. Pairwise repertoire similarity was assessed using repOverlap, which was used to compute the Jaccard, Tversky, Cosine, and Morisita overlap indices. Shared clonotypes across samples were identified using trackClonotypes. In this study, the term “clonotype” refers, unless otherwise specified, to the unique amino acid (aa) sequence of the CDR3 region. The overlap of CDR3aa sequences between TILs and PBMCs, or among different TILs, was analyzed using the “Repertoire Overlap” function from the Immunarch package, while shared CDR3aa sequences among TILs were identified with “Track Clonotype”. The number of clonotypes was estimated using the repExplore function.

The TCR repertoire NGS raw data generated and analyzed during this study have been deposited in the Sequence Read Archive (SRA) under Project ID PRJNA1459400. The project contains 20 datasets, each one identified by a sequential accession number from SRX33156384 to SRX33156403. Below, we provide the direct access links (URLs) for each dataset included in the study:

Accession number SRX33156384: https://www.ncbi.nlm.nih.gov/sra/SRX33156384[accn];

Accession number SRX33156385: https://www.ncbi.nlm.nih.gov/sra/SRX33156385[accn];

Accession number SRX33156386: https://www.ncbi.nlm.nih.gov/sra/SRX33156386[accn];

Accession number SRX33156387: https://www.ncbi.nlm.nih.gov/sra/SRX33156387[accn];

Accession number SRX33156388: https://www.ncbi.nlm.nih.gov/sra/SRX33156388[accn];

Accession number SRX33156389: https://www.ncbi.nlm.nih.gov/sra/SRX33156389[accn];

Accession number SRX33156390: https://www.ncbi.nlm.nih.gov/sra/SRX33156390[accn];

Accession number SRX33156391: https://www.ncbi.nlm.nih.gov/sra/SRX33156391[accn];

Accession number SRX33156392: https://www.ncbi.nlm.nih.gov/sra/SRX33156392[accn];

Accession number SRX33156393: https://www.ncbi.nlm.nih.gov/sra/SRX33156393[accn];

Accession number SRX33156394: https://www.ncbi.nlm.nih.gov/sra/SRX33156394[accn];

Accession number SRX33156395: https://www.ncbi.nlm.nih.gov/sra/SRX33156395[accn];

Accession number SRX33156396: https://www.ncbi.nlm.nih.gov/sra/SRX33156396[accn];

Accession number SRX33156397: https://www.ncbi.nlm.nih.gov/sra/SRX33156397[accn];

Accession number SRX33156398: https://www.ncbi.nlm.nih.gov/sra/SRX33156398[accn];

Accession number SRX33156399: https://www.ncbi.nlm.nih.gov/sra/SRX33156399[accn];

Accession number SRX33156400: https://www.ncbi.nlm.nih.gov/sra/SRX33156400[accn];

Accession number SRX33156401: https://www.ncbi.nlm.nih.gov/sra/SRX33156401[accn];

Accession number SRX33156402: https://www.ncbi.nlm.nih.gov/sra/SRX33156402[accn];

Accession number SRX33156403: https://www.ncbi.nlm.nih.gov/sra/SRX33156403[accn].

Large, processed data tables generated using MiXCR, are available at Zenodo (https://doi.org/10.5281/zenodo.17733884). Other data supporting the findings of this study are included in the article and its supplementary materials.

For single-cell analysis, approximately 2 × 10⁶ cells were cryopreserved and shipped on dry ice to Genewiz, where single-cell RNA sequencing libraries were prepared using the 10x Genomics® Chromium™ platform and sequenced on an Illumina® NovaSeq™ 6000 instrument. Following quality control, a mean of 1,971 T cells per sample (range: 1,128–2,489) were retained for downstream analyses. Bioinformatic analysis of the receptor transcripts revealed a high efficiency of TRA/TRB pair reconstruction, with an average of 80.4% of cells showing productive pairing of both chains (range: 72.4–85.9%). Moreover, 73.5% of the analyzed T cells displayed a single, productive, and unique α/β clonotypic pair (range: 66.9–80.4%).

### TCR repertoire diversity analysis

To address TCR repertoire diversity we calculated multiple clonality indices. Richness is defined as the total number of unique clonotypes without considering their relative abundances. To account for differences in sequencing depth, normalized richness was calculated by dividing TCR richness with the number of total productive TCR nucleotide sequences. Shannon diversity provides a comprehensive measure of repertoire heterogeneity by integrating both clonotype richness and evenness. The Shannon entropy reflects both the diversity and distribution of clonotypes within a repertoire [42], [43]. The Pielou’s Evenness Index evaluates how uniformly clonotypes are distributed across the repertoire. The Simpson index measures the probability that two randomly chosen individuals belong to the same clonotype and gives more weight to the most abundant clonotypes (dominance).

To quantify repertoire overlap we used the Jaccard index, which assesses similarity by dividing the size of the intersection (shared clonotypes) by the size of the union (total distinct clonotypes) of the two repertoires [43], [44] . For cases where an asymmetric similarity measure is needed, the Tversky index offers an alternative [45]. The Cosine similarity takes a vector-based approach to measure the similarity between two non-zero distributions of clonotypes [46] . The Morisita’s overlap index is commonly used in ecology to assess population dispersion but is equally applicable to immune repertoire analysis as it measure considers the abundance of clonotypes [43], [47] . For a comprehensive discussion of the theoretical basis and immunological applications, readers are refered to the review by Aversa et al., and Chifelle et al.[43], [48].

### In-silico analysis

Clustering of CDR3β amino acid sequences was performed using the GLIPH2 algorithm from the turboGLIPH R package, a computational framework designed to analyze TCR repertoires by clustering TCR sequences into specificity groups. For the analysis of the most abundant intratumoral clonotypes, the query set consisted of the ten CDR3β in the biopsy repertoire with the highest frequency. For the analysis of the total intratumoral clonotypes, the query set consisted of all the intratumoral CDR3β clonotypes across all patients, and CDR3β sequences were excluded only when not detected in the matched PBMC dataset. The reference set comprised previously reported CDR3 amino acid sequences associated with known epitopes, obtained from the IEDB [49], and was used as a background for enrichment analysis. For each specificity group, a contingency table comparing the frequency of CDR3β sequences in the query and reference sets was constructed, and statistical enrichment was assessed using Fisher’s exact test. Clusters containing fewer than 3 (for the ten most abundant intratumoral clonotype analysis) or 10 (for the analysis of intratumoral clonotypes across patients) CDR3β sequences were excluded from downstream analyses. Local similarity groups were defined by shared short CDR3β motifs, consistent with antigen-driven selection, whereas global similarity groups were defined by broader sequence homology across the CDR3β region. Statistical significance of specificity groups was evaluated using Fisher’s exact test p-values, with lower p-values indicating higher confidence in non-random convergence [50]. In addition, the probability of interaction between CDR3aa sequences and peptides was evaluated using the ERGO-II tool [51].

### Antigens

The peptides restricted to the most frequent class I and II HLA alleles in the Caucasian population (HLA-A*01:01; A*02:01; A*03:01; B*07:02; B*15:01; B*08:01; DRB1*01:01; DRB1*01:04), derived from PAP (UniProt ID P15309), PSCA (UniProt ID O43653), TGM4 (UniProt ID P49221), PSA (UniProt ID P07288), PSMA (GenBank ID AAM34479), AMACR (UniProt ID Q9UHK6) and PSGR (GenBank ID AAK38728), were identified using the NetMHCpan tool (v 4.0 and 3.2) [52], [53]. Peptides were then purchased in lyophilized form (GenScript Biotech Company, Nanjing, China) and reconstituted in either dimethyl sulfoxide (DMSO) or water, depending on their hydrophobic properties, at a concentration of 40 mg/mL. For each antigen, a peptide pool was prepared by diluting the individual peptides in water, to be used in PBMCs expansion and functional assays.

### T-cell expansion and Enzyme-linked ImmunoSpot Assay

For in vitro T cell expansion, fresh PBMCs were isolated from whole blood samples by density gradient centrifugation and immediately cultured in replicates of 2–3 × 10⁵ cells/well in 96-well U-bottom plate in AIM-V medium, 50 μg/mL streptomycin, 10 μg/mL gentamicin sulfate (code 12055091; LIFE Technologies), supplemented with 5% human AB serum (code H3667; Sigma-Aldrich), 10 U/mL IL-2 (code 200-02; ThermoFisher Scientific), 20 µM β-mercaptoethanol (code M3148; Sigma-Aldrich,), and 1 mM HEPES (code S-H0887; Sigma-Aldrich). The cells were then stimulated for 15 days with 2 μg/mL of each antigen pool, with IL-2 addition and medium refreshment every 2–3 days. T cell responses before and after expansion was evaluated using the Enzyme-linked ImmunoSpot (ELISPOT) assay (cat. 3420-4AST-P1-1, Mabtech, Nacka Strand, Sweden) in response to the antigen pool at 2 μg/mL or the polyclonal activator anti-CD3 supplied with the kit, according to the manufacturer’s instructions. Spots corresponding to stimulated cells secreting IFNγ were counted. The ImmunoSpot Forming Units (SFU) were then quantified using the ELISpot IRIS2 Plate Reader (Mabtech). The IFNγ-ELISPOT data were reported as stimulating forming units × 10⁶ PBMCs (SFU/10⁶), calculated for each PBMC sample by subtracting the spots from the unstimulated wells from the spots of the peptide-stimulated wells and normalizing to 10⁶ PBMCs [54], or compared to the polyclonal activator anti-CD3. The T cell expansions were considered significant when the percentage of spots obtained with the antigen pool, relative to those obtained with anti-CD3, exceeded 1%, indicating a measurable T cell response.

### Statistical analysis

Statistical tests were selected based on appropriate assumptions with respect to data distribution and variance characteristics. The Fisher’s exact test was used to compare prevalence between groups, the Mann-Whitney test was applied to compare unpaired continuous data not normally distributed, and the Wilcoxon matched-pairs signed rank test was used as a nonparametric method to compare matched data; p values < 0.05 were considered statistically significant. Pairwise TIL–TIL repertoire overlap was analyzed using a patient-level summary approach to account for the non-independence of pairwise comparisons. For each patient, the mean overlap with all other patients within the same disease-grade group was calculated, yielding one independent observation per subject. Differences between low- and high-grade tumors were assessed using an exact permutation test based on all possible allocations of four low-grade and six high-grade patients (210 permutations). Effect sizes were quantified using rank-biserial correlation and Cliff’s delta with 95% bootstrap confidence intervals (20,000 iterations). Statistical significances are reported in the figures and/or the figure legends. Statistical tests were performed with GraphPad PRISM software 9.3 (GraphPad Software, La Jolla, CA, USA) or R (version 4.2.3) [55] .

## RESULTS

### Sample eligibility for TCRβ repertoire analysis

Total RNA was extracted from 26 tumor biopsies and evaluated for quality and integrity. Eight samples were excluded due to insufficient RNA yield, most likely reflecting the limited amount of processed tissue. Of the remaining 18 biopsies, 10 generated functional TCRβ libraries suitable for NGS, whereas 8 failed library generation, indicating either very low levels of T-cell-derived RNA or RNA levels below the analytical sensitivity required for successful TCRβ library generation.

The included and excluded patients showed comparable baseline clinical characteristics (Table 1). No obvious differences were observed between the two groups with respect to PSA (10±8.9 vs 20.3±17.3 ng/mL), histopathological diagnosis (predominantly acinar adenocarcinoma), Gleason score (predominantly 7:3+4), Grade Group (1.7/5 vs 1.5/5), or major comorbidities (predominantly hypertension). While we cannot completely exclude the possibility that successful library generation may be associated with biological factors, these data suggest that the excluded samples presented low T-cell infiltration.

**Table 1:** Clinical Informations of the Study Participants.

| Patient ID | Gender | Total PSA (ng/ml) | Istologic al Classific | Gleason Score | Grade Group | Co-morbidities | Included/ Excluded |
| --- | --- | --- | --- | --- | --- | --- | --- |
|  |  | ) | ation |  |  |  |  |
| 1193 | Male | 7 | Acinar<br>Adenocarcinoma | 6 | 1/5 | Dyslipidemia | Included |
| 1194 | Male | 20 | Acinar<br>Adenocarcinoma | 8:4+4 | 4/5 | Hypertension | Included |
| 1262 | Male | 9.5 | Acinar<br>Adenocarcinoma | 8:4+4 | 4/5 | Hypertension,<br>dyslipidemia | Included |
| 1268 | Male | 7.49 | High<br>Grade<br>PIN | N/A | N/A | None | Included |
| 1354 | Male | 7.48 | Acinar<br>Adenocarcinoma | 7:3+4 | 2/5 | Hypothyroidism due to<br>cancer | Included |
| 1517 | Male | 16.27 | Acinar<br>Adenocarcinoma | 7:3+4 | 2/5 | None | Included |
| 1543 | Male | 13.9 | High<br>Grade<br>PIN | N/A | N/A | Hypertension | Included |
| 1544 | Male | 3 | Acinar<br>Adenocarcinoma | 7:3+4 | 2.5 | Heart failure,<br>mito-aortic<br>valvular<br>disease,<br>osteoarthritis<br>and<br>osteoporosis | Included |
| 1572 | Male | 9.36 | Acinar<br>Adenocarcinoma | 7:3+4 | 2/5 | Hypertension | Included |
| 1834 | Male | 27.49 | Intraductal<br>Carcinoma | N/A | N/A | Hypertension,<br>dyslipidemia | Included |
| 1195 | Male | 2.59 | High<br>Grade<br>PIN | N/A | N/A | Hypertension | Excluded |
| 1260 | Male | 10.2 | Acinar<br>Adenocarcinoma | 6 | 1/5 | Hypertension | Excluded |
| 1264 | Male | 4.34 | Acinar<br>Adenocarcinoma | 6 | 1/5 | Hypertension,<br>low-grade<br>non-invasive<br>urothelial<br>carcinoma | Excluded |
| 1269 | Male | 36 | Acinar<br>Adenocarcinoma | 7:4+3 | 3/5 | Hypertension | Excluded |
| 1291 | Male | 41 | Acinar<br>Adenocarcinoma | 7:4+3 | 3/5 | Hypertension | Excluded |
| 1292 | Male | 5.68 | Acinar<br>Adenocarcinoma | 6 | 1/5 | Hypertension,<br>dyslipidemia | Excluded |
| 1296 | Male | 4.14 | High<br>Grade<br>PIN | N/A | N/A | Thyroid<br>carcinoma | Excluded |
| 1347 | Male | 12.99 | Acinar<br>Adenocarcinoma | 7:3+4 | 2/5 | None | Excluded |
| 1353 | Male | 4.6 | Acinar<br>Adenocarcinoma | 7:4+3 | 3/5 | Hypertension | Excluded |
| 1489 | Male | 6.27 | Acinar<br>Adenocarcinoma | 6 | 1/5 | None | Excluded |
| 1491 | Male | 4.98 | Acinar<br>Adenocarcinoma | 6 | 1/5 | Dyslipidemia,<br>prediabetes | Excluded |
| 1538 | Male | 15.91 | Acinar<br>Adenocarcinoma | 7:3+4 | 2/5 | Hypertension,<br>dyslipidemia | Excluded |
| 1540 | Male | 15 | Acinar<br>Adenocarcinoma | N/A | N/A | Hypertension,<br>dyslipidemia | Excluded |
| 1544 | Male | 3 | Acinar<br>Adenocarcinoma | 6 | 1/5 | Heart failure | Excluded |
| 1575 | Male | 4.47 | Acinar<br>Adenocarcinoma | 7:3+4 | 2/5 | Hypertension | Excluded |
| 1623 | Male | 6.47 | Acinar<br>Adenocarcinoma | 7:4+3 | 3/5 | Hypertension | Excluded |

For the same 10 patients, TCRβ libraries were also successfully prepared from PBMCs isolated from residual diagnostic samples. Collectively, sequencing yielded 98,387 distinct clonotypes (unique CDR3β amino acid sequences) from TIL repertoires and 312,500 clonotypes from PBMC repertoires (Table 2 and Supplementary Data).

**Table 2:** Sequencing Information.

| **Table 2: Sequencing Information** |  |  |  |  |  |  |
| --- | --- | --- | --- | --- | --- | --- |
| **Patient ID** | **n_sequence_biopsy** | **n_sequence_PBMCs** | **n_cln\* biopsy** | **n_cln\* PBMCs** | **n_unique_cln\*\* biopsy** | **n_unique_cln\*\* PBMCs** |
| 1193 | 2680156 | 636112 | 17778 | 23537 | 5361 | 17964 |
| 1194 | 2583724 | 7530972 | 26378 | 95481 | 9804 | 34007 |
| 1262 | 1500484 | 1206768 | 26919 | 55334 | 11583 | 40316 |
| 1268 | 1051234 | 676880 | 13602 | 21610 | 5373 | 16769 |
| 1354 | 2463604 | 915392 | 15992 | 45155 | 4844 | 34332 |
| 1517 | 2004990 | 978086 | 34773 | 20619 | 15516 | 16710 |
| 1543 | 22922856 | 417900 | 21093 | 26409 | 6807 | 22306 |
| 1544 | 3316748 | 562272 | 26054 | 55451 | 6317 | 44378 |
| 1572 | 3120026 | 959932 | 36012 | 75626 | 17738 | 60232 |
| 1834 | 1773228 | 404122 | 34209 | 29013 | 15044 | 25486 |

\* = n\_cln - number of unique CDR3β nucleotide sequences
\*\* = n\_unique\_cln—number of unique CDR3β amino acid sequences

Overall, only 10 of the 26 diagnostic biopsies yielded successful TCRβ libraries, indicating that detectable T-cell-derived RNA was present in only a subset of samples and generally at low abundance, consistent with the sparse T-cell infiltration typically observed in the immunologically "cold" prostate tumor microenvironment [26].

### TCRβ repertoires of TILs display a distinct clonotype distribution compared with peripheral blood T cells

We initially compared the overall clonality and diversity of tumor and peripheral blood TCRβ repertoires using commonly employed repertoire metrics [48] . Peripheral repertoires exhibited significantly higher richness, indicating a greater number of unique clonotypes compared to TIL repertoires, despite comparable sequencing depth (Fig. 1A and B). No significant differences were observed in repertoire metrics related to the evenness of the clonotype distribution or in those sensitive to rare clonotype species, including Shannon entropy, Shannon diversity, and Pielou’s evenness (Fig. 1C–E). By contrast, the Simpson index was lower in TIL repertoires, indicating reduced evenness and a greater dominance of expanded clonotypes within the tumor microenvironment (Fig. 1F).

**Figure 1:**
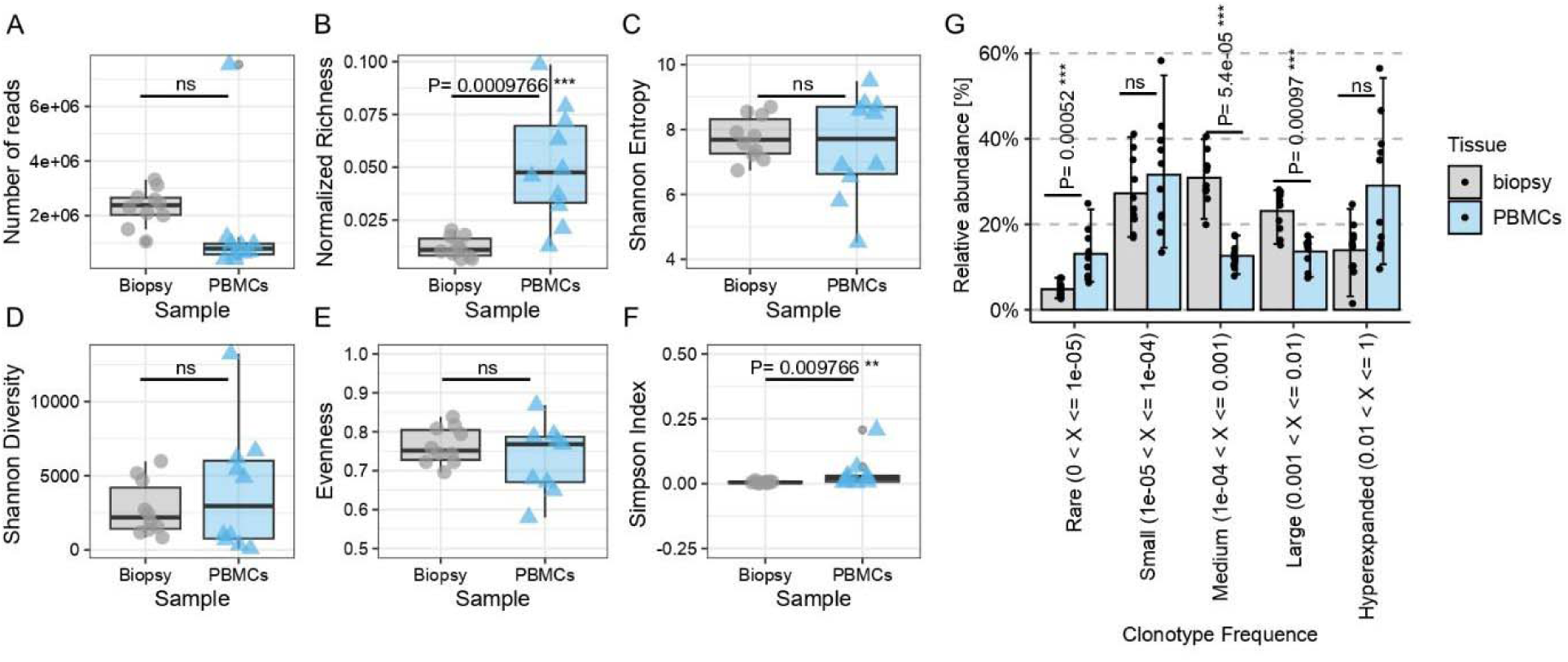
Comparison of peripheral T cells or TILs (Biopsy) repertoire metrics. **A** number of total reads. **B** richness value normalized by the number of total sequence reads. **C** Shannon entropy. **D** Shannon diversity. **E** Pielou’s Eveness. **F** Simpson Index. **G** relative abundance (%) of clonotypes based on clonotype frequence grouped in rare, small, medium, large and hyperexpanded. Statistical comparisons between groups were performed using the Wilcoxon rank-sum test.

To evaluate the potential influence of sequencing depth on repertoire diversity, we examined the correlation between sequencing reads and each diversity metric (Supplementary Fig. 2). As expected, richness showed a modest positive association with sequencing depth (Pearson *r* = 0.450), whereas Shannon entropy, Shannon diversity, Pielou’s evenness, and the Simpson index showed only weak correlations. Because normalized richness is mathematically derived from sequencing depth, its correlation with read count was not evaluated. Overall, these analyses indicate that sequencing depth had minimal influence on the diversity metrics used in this study. Accordingly, all analyses were performed using the complete repertoires without rarefaction or downsampling in order to preserve sequencing information.

To further characterize the clonotype distribution within the repertoires, we binned unique clonotypes based on their abundance as rare, small, medium, large, and hyperexpanded. A greater fraction of the repertoires derived from biopsies was occupied by medium and large clonotypes compared to the repertoires derived from PBMCs, indicating a repertoire architecture enriched for high-frequency clonotypes and reduced representation of low-frequency clonotypes. (Fig. 1G).

### Analysis of repertoire overlap suggests distinct patterns associated with cancer severity

We characterized the overlap between TILs and PBMCs repertoires, by quantifying the fraction of clonotypes shared between the two compartments (Fig. 2A). Patients were grouped according to Grade Group (Grade Group 1 versus Grade Group ≥2). The number of shared clonotypes did not differ significantly between the two groups (Fig. 2B). Effect size analysis indicated a negligible difference (rank-biserial correlation, rRB = 0.00, 95% CI −0.64 to 0.64; Cliff’s δ = 0.00, 95% CI −0.72 to 0.72), although the confidence intervals were wide, reflecting the limited statistical power of this exploratory cohort.

**Figure 2:**
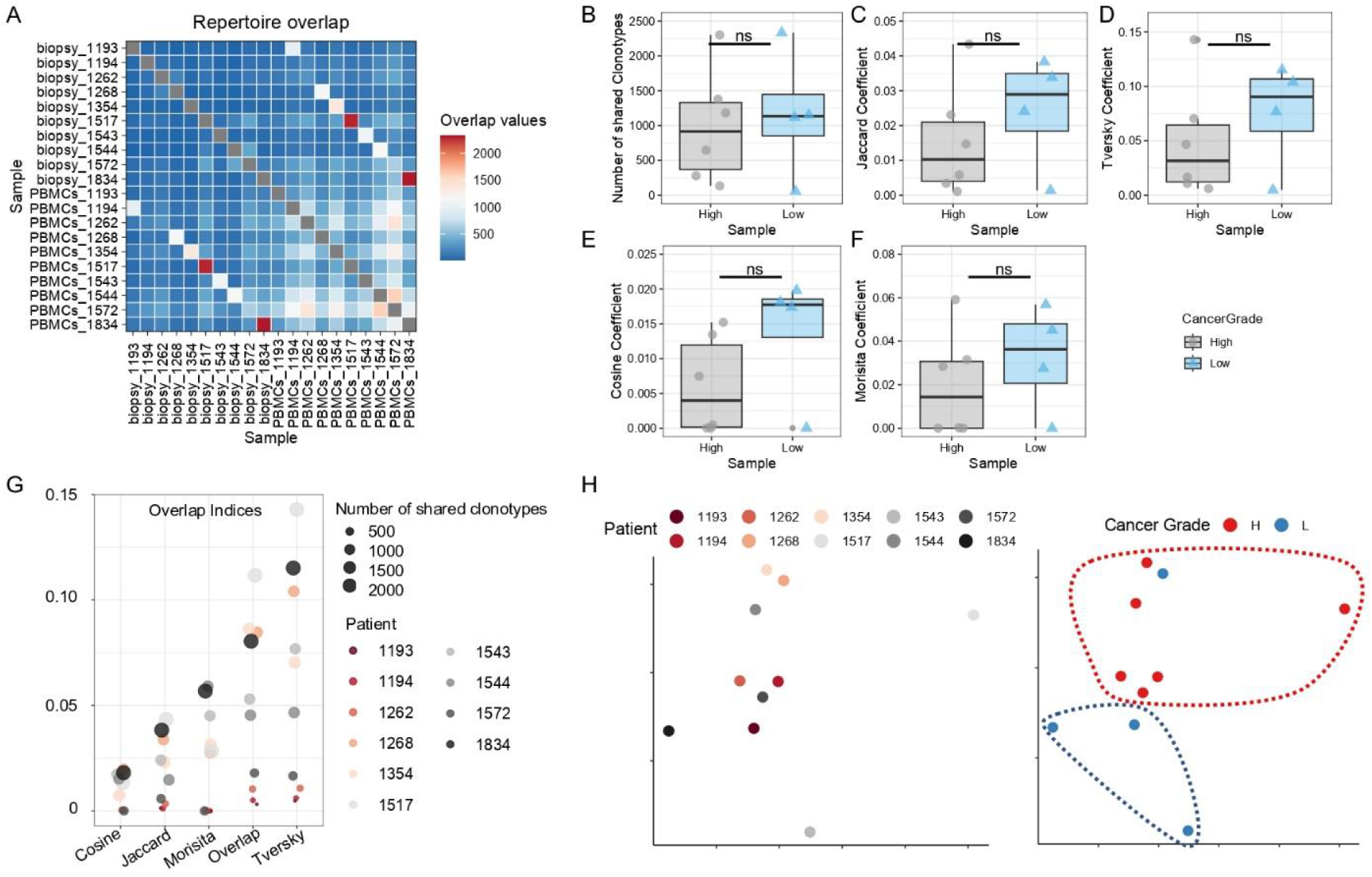
Analysis of repertoire overlap in high- and low-ISUP cancer grade samples. **A** matrix representation of overlap values between PBMC and matched tumor biopsy repertoires for each patient. **B**, **C**, **D**, **E** and **F** Number of shared clonotypes (Overlap), Jaccard, Tversky, Cosine, and Morisita coefficients. Statistical comparisons between grade groups were performed using the Wilcoxon rank-sum test, and effect sizes were quantified using rank-biserial correlation and Cliff’s delta with 95% confidence intervals. **G** Bubble chart showing overlap coefficient values color-coded by patient, and with size scaled by the number of shared clonotypes. **H** tSNE visualization based on the combined overlap metrics. Samples are color-coded according to patient identity or cancer grade. The t-SNE embedding is shown for exploratory visualization of repertoire similarity patterns.

To further assess repertoire overlap, we calculated complementary frequency-aware overlap metrics, including the Jaccard coefficient, Tversky index, Cosine similarity, and Morisita overlap index (Fig. 2C–G). None of these measures differed significantly between Grade Group 1 and Grade Group ≥2 tumors. However, the Jaccard coefficient (rRB = 0.33), Tversky index (rRB = 0.25), and Cosine similarity (rRB = 0.50) showed small-to-moderate effect sizes consistent with a tendency toward greater repertoire overlap in Grade Group 1 tumors, whereas Morisita overlap showed only a negligible effect (rRB = 0.08). Confidence intervals were broad for all estimates, indicating substantial uncertainty due to the limited sample size. Accordingly, these findings should be regarded as exploratory and require validation in larger cohorts.

Finally, we applied t-distributed stochastic neighbor embedding (t-SNE) to visualize the overall similarity among samples using all overlap metrics simultaneously (Fig. 2H). The resulting embedding suggested a tendency for samples to cluster according to Grade Group, with most Grade Group 1 tumors occupying a common region of the projection. Because t-SNE is an unsupervised visualization method and does not provide formal statistical evidence of group separation, this observation should be interpreted cautiously. Overall, these analyses suggest that integrating multiple complementary overlap metrics may reveal repertoire relationships that are not captured by individual indices alone.

### Pairwise TILs repertoire overlap reveals increased clonotype shared in patients with more severe cancer

To investigate whether intratumoral repertoire similarity differed according to disease severity, we compared pairwise overlap among TIL repertoires after summarizing pairwise comparisons at the patient level to account for their non-independence. Grade Group ≥2 tumors showed greater repertoire similarity than Grade Group 1 tumors when assessed by the number of shared clonotypes (Wilcoxon rank-sum test, *P* = 0.0095), the Jaccard coefficient (*P* = 0.019), and the Tversky index (*P* = 0.019) (Fig. 3A–C). Corresponding effect size estimates were large (Supplementary Table 2). In contrast, Cosine similarity and Morisita overlap did not differ significantly between groups (Fig. 3D,E). Differences between low- and high-grade tumors were further evaluated using an exact permutation test based on all 210 possible assignments preserving the observed group sizes. The revised analysis showed a significantly greater number of shared clonotypes among high-grade tumors (P = 0.043). Jaccard and Tversky indices showed large effect-size estimates and trends toward greater similarity in high-grade tumors, although they did not reach the conventional significance threshold in the exact permutation analysis (P = 0.052 for both). Cosine similarity and Morisita overlap were not significantly different between groups (Supplementary Table 2).

**Figure 3:**
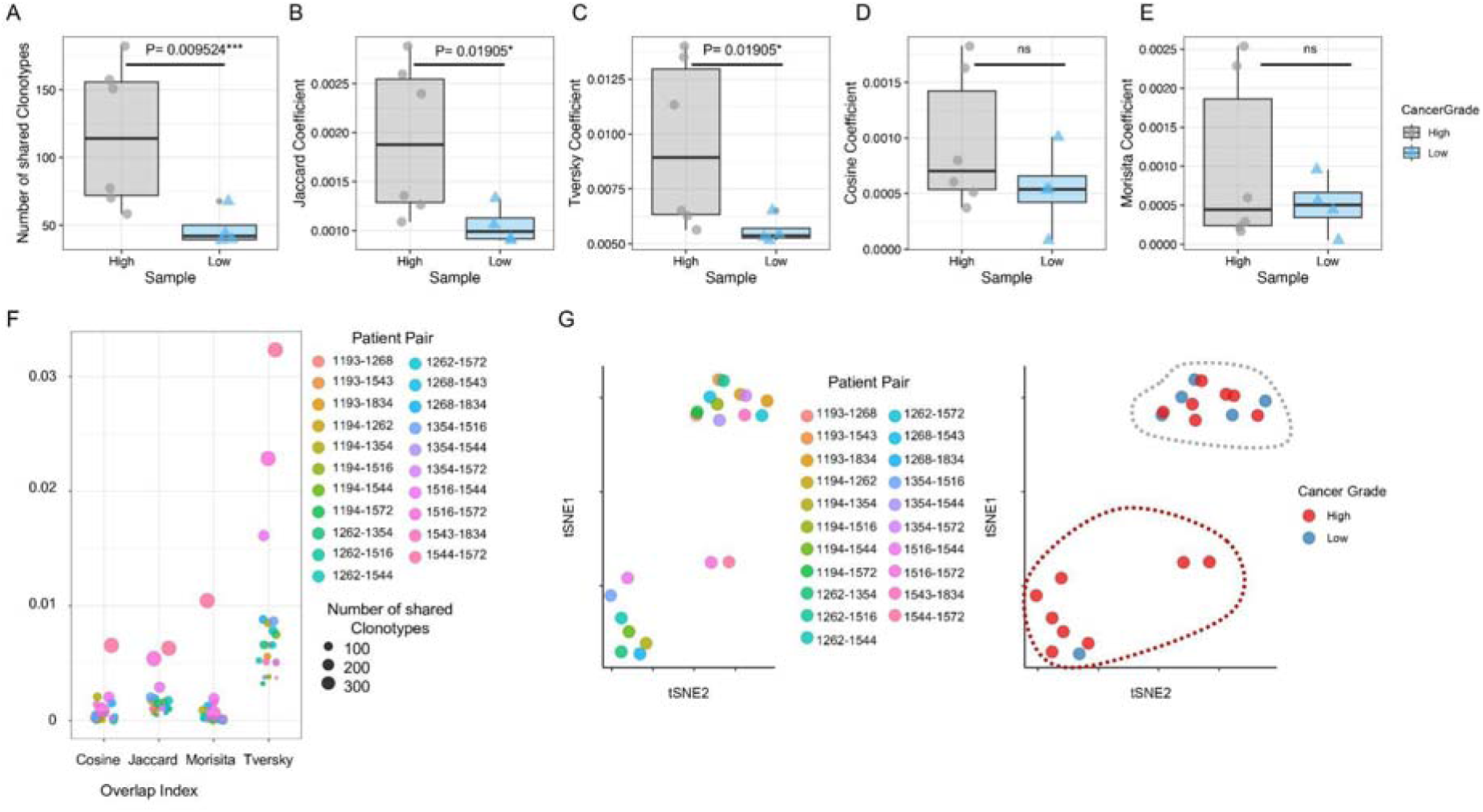
Pairwise overlap of repertoires derived from tumor biopsies in high- and low-cancer grade. **A, B, C, D**, and **E** Comparison of biopsy repertoire similarity between high- and low-grade tumors based on (A) number of shared clonotypes, (B) Jaccard coefficient, (C) Tversky coefficient, (D) Cosine similarity, and (E) Morisita overlap index. For statistical comparisons, pairwise overlap values were summarized at the patient level by calculating the mean overlap of each biopsy with all other biopsies within the same grade group, resulting in one independent value per patient. Differences between groups were assessed using the Wilcoxon rank-sum test, and effect sizes were estimated using rank-biserial correlation and Cliff’s delta with 95% confidence intervals. **F** Bubble chart showing Cosine, Jaccard, Morisita, and Tversky coefficient values. Each point represents a unique patient–patient comparison, with colors indicating patient pairs and point size proportional to the number of shared clonotypes. These pairwise values were not treated as independent observations for statistical testing. **H** tSNE clustering plot, color coded by patient pair or cancer grade.

Individual pairwise values for each index are presented in Fig. 3F, as an exploratory visualization of repertoire similarity patterns among biopsy samples. Each point represents a unique patient–patient comparison, with point color indicating the patient pair and point size representing the absolute number of shared clonotypes. We further applied t-distributed stochastic neighbor embedding (t-SNE) using the complete set of overlap metrics (Fig. 3G). The resulting embedding suggested a tendency for Grade Group ≥2 samples to cluster more closely than Grade Group 1 samples. However, because t-SNE is an unsupervised visualization method, this observation should be considered exploratory and not as formal evidence of group separation. Overall, these findings are consistent with greater inter-patient repertoire convergence among Grade Group ≥2 tumors, although the biological mechanisms underlying this convergence remain to be established.

### Insights into most abundant TILs clonotypes specificity

To get a better understanding of the sequence determinants of intratumoral clonotypes, we focused our analysis on the most abundant species within the tumor biopsy repertoires. On average, we observed that only 3.6 ± 3.7 of the 10 most abundant TIL-derived clonotypes were also detected in the matched PBMC repertoires, indicating a variable degree of systemic overlap. Therefore, we narrowed our analysis to the shared portion of the repertoires. The top 10 intratumoral clonotypes and their relative frequencies compared to the PBMC counterpart are shown in Fig. 4A, showing that they are generally more abundant within the TILs repertoires compared to the peripheral repertoires. Notably, there was no sequence overlap in these top clonotypes across the ten patients.

**Figure 4:**
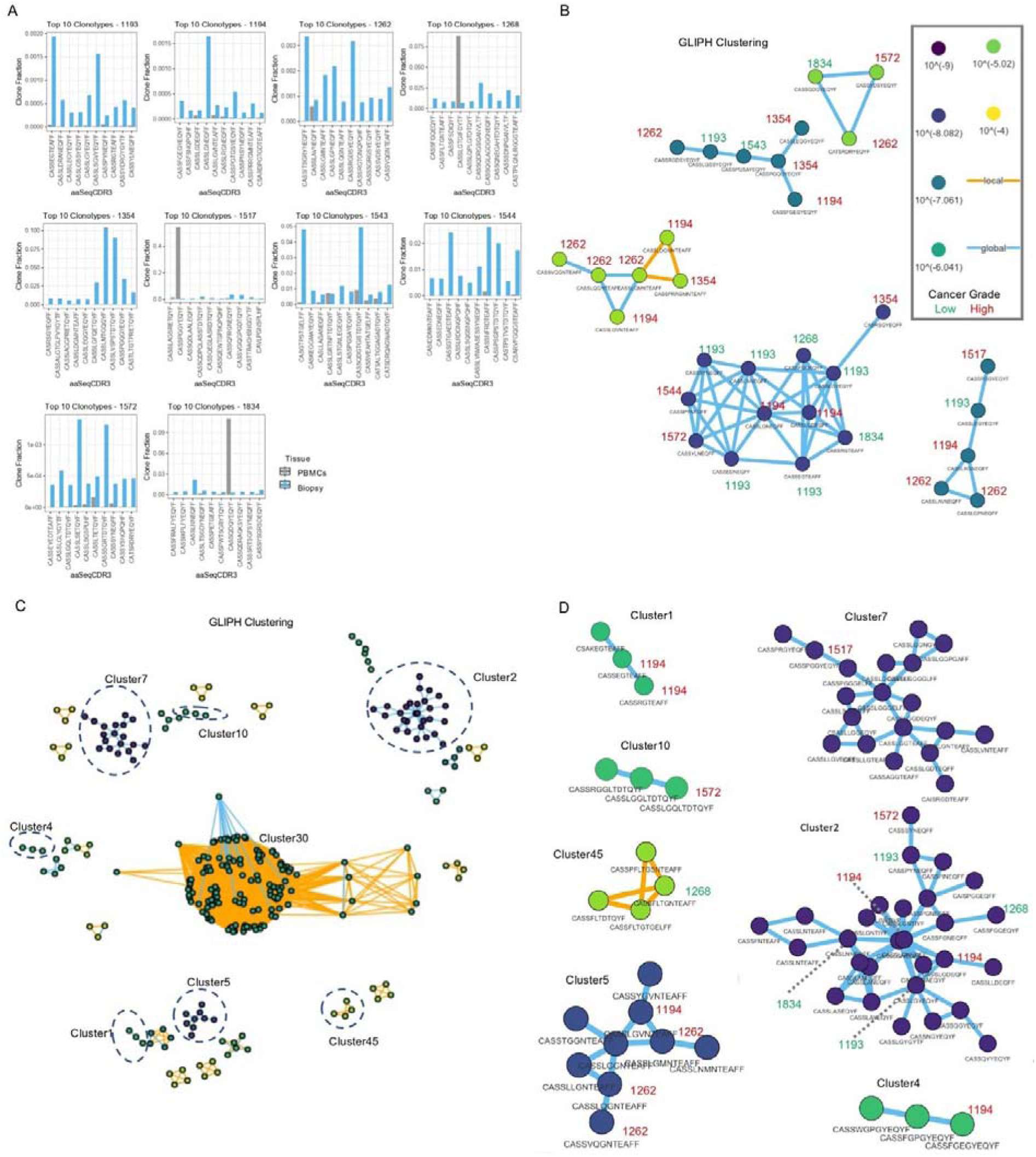
Analysis of the top ten most frequent clonotypes within TILs. **A,** Barplots showing clonotype frequency and amino acidic sequence of the most abundant intratumoral clonotypes per patient compared to their frenquency within peripheral repertoires. **B,** GLIPH2 clustering of the top ten most frequent intratumoral clonotypes. **C,** GLIPH2 clustering of intratumoral clonotypes with putative TAA-binding clonotypes from the IEDB database. Dotted blue circled highlight clusters containing both intratumoral clonotypes and known TAA-binding clonotypes. **D** Clusters containing both clonotypes from IEDB and intratumoral clonotypes from our dataset. Clonotype sequence and Patient ID numbers are reported.

To investigate whether these distinct clonotypes might nevertheless share common antigen-recognition features, we applied the Grouping of Lymphocyte Interactions by Paratope Hotspot 2 (GLIPH2) algorithm, a computational framework designed to cluster TCR sequences into putative specificity groups[50] . Analysis of the top 10 intratumoral clonotypes identified five GLIPH2 clusters, suggesting the presence of conserved CDR3β sequence motifs (Fig. 4B). We then extended the analysis by including TCR sequences annotated in the IEDB as recognizing tumor-associated antigens (Fig. 4C). GLIPH2 identified seven clusters containing both intratumoral clonotypes and TAA-associated reference sequences (Fig. 4D). These clusters included clonotypes annotated in association with several well-established cancer-related antigens, including NY-ESO-1 (CTAG1B), Wilms’ tumor protein (WT1), TP53, CCND1, KAT6A (MOZ), and PMEL (gp100) (Table 3). Although these computational analyses do not establish direct TCR–peptide–HLA specificity, they suggest that highly expanded intratumoral clonotypes may share structural features with previously described tumor-associated TCRs and provide a rationale for subsequent functional validation.Characterization of intratumoral clonotype specificity

**Table 3:**
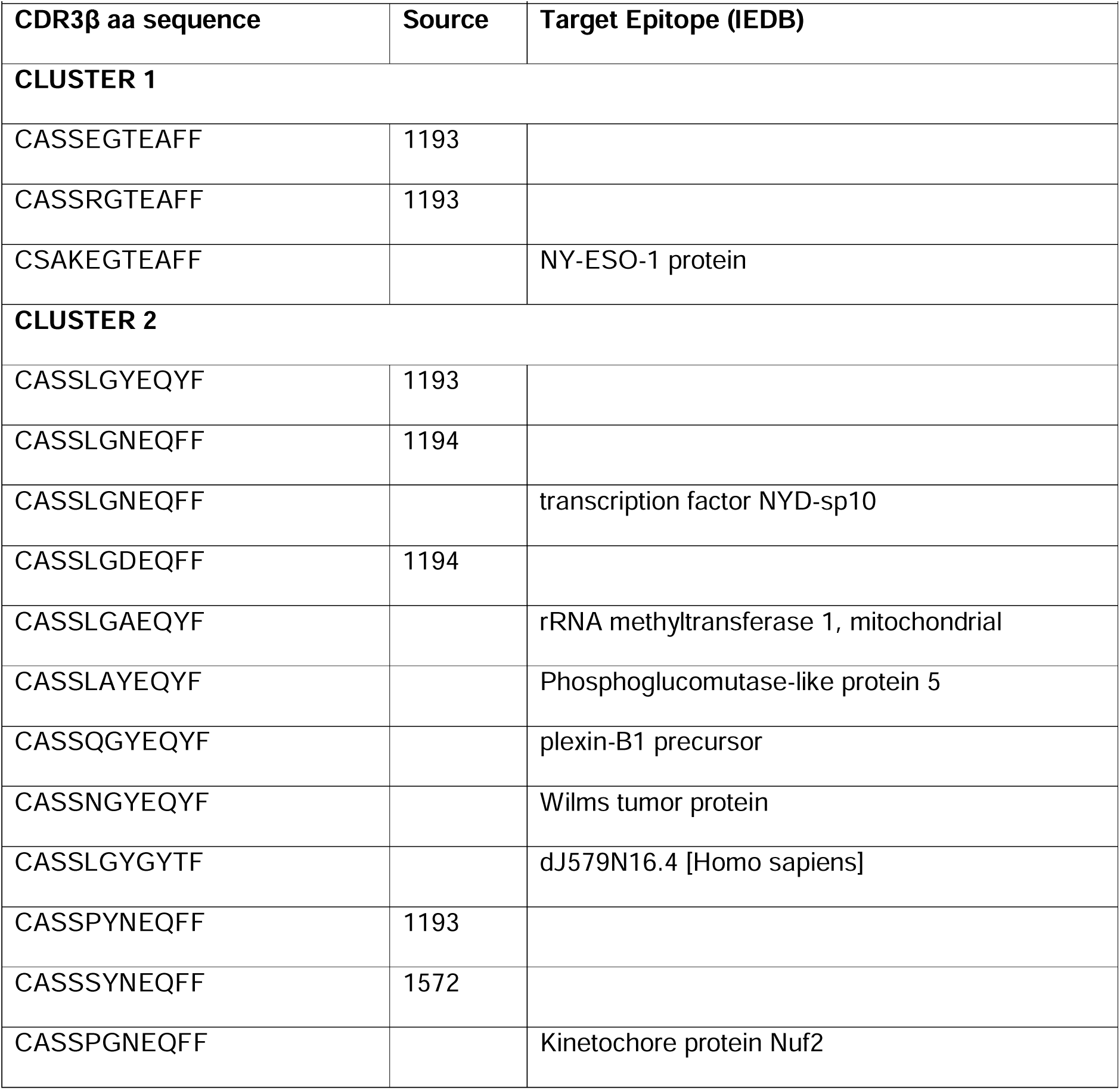
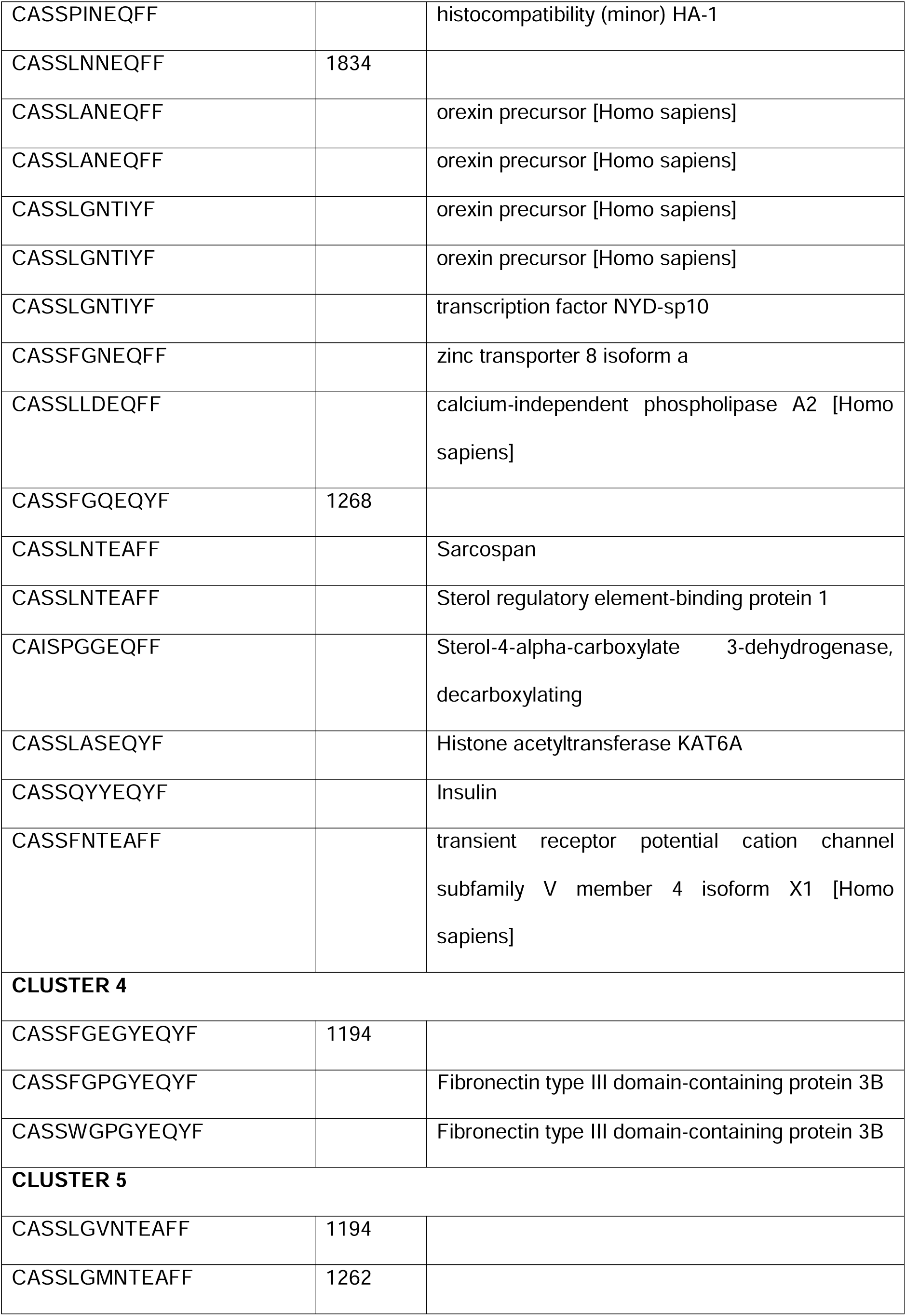
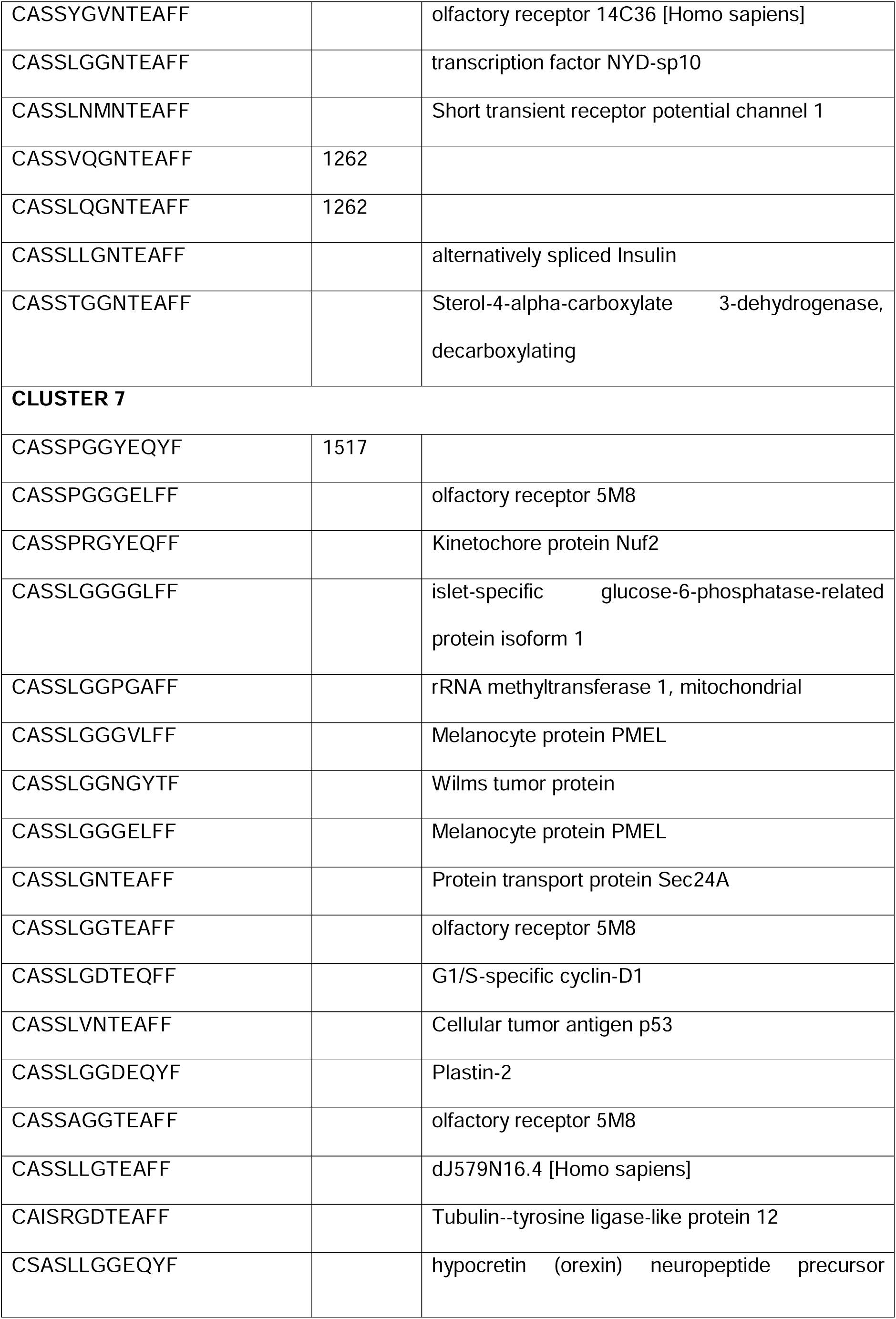
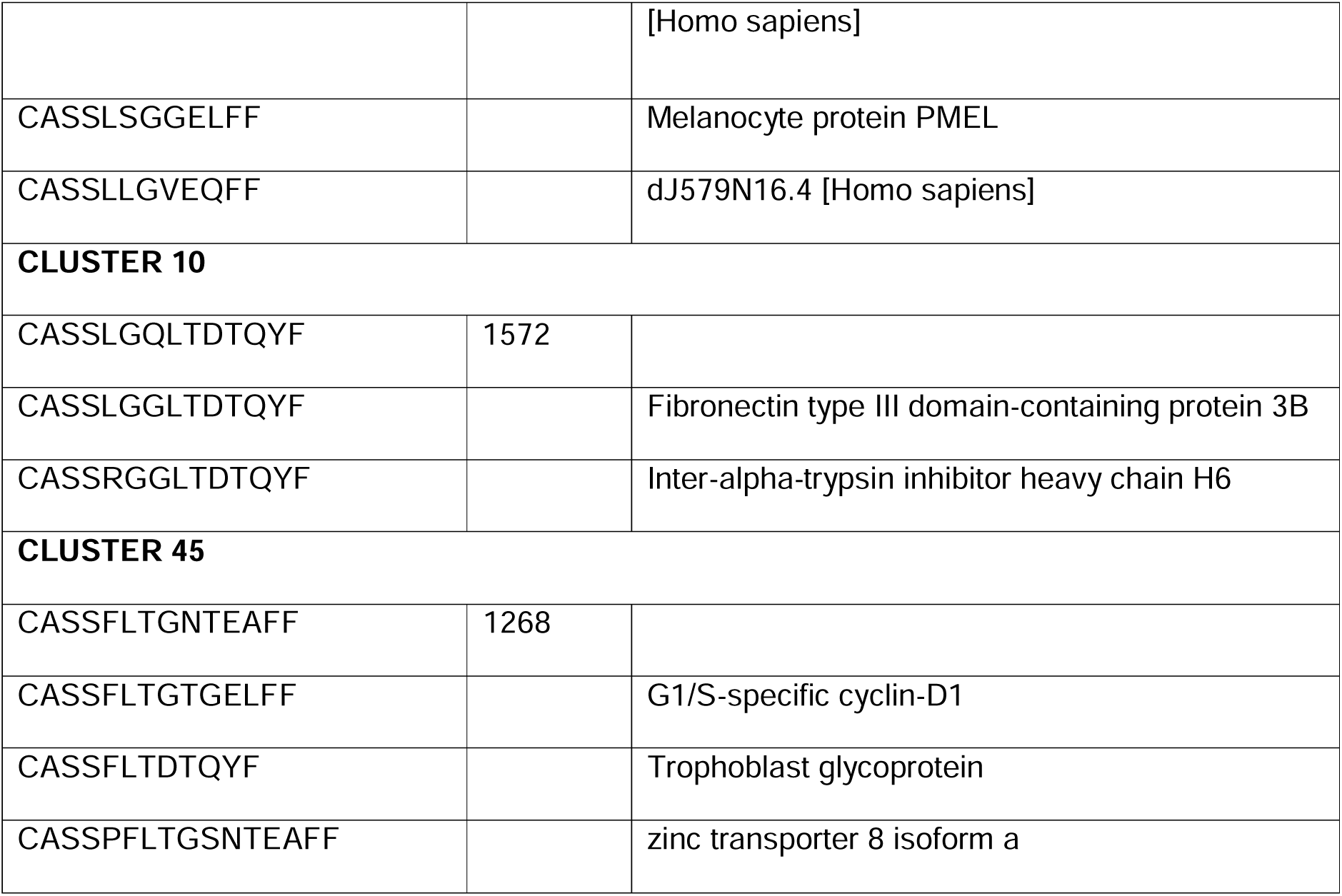
Intratumoral Clonotype Clusters.

The characterization of TILs clonotype sequences is crucial to identify molecular players that can be targeted by novel immunotherapies. For this reason, we generated a repertoire overlap matrix and evaluated clonotype sharing across patients, revealing a range of 20 to 389 intratumoral clonotypes shared within patients (Fig. 5A). With the aim of getting insights into epitope specificity of these intratumoral clonotypes, we applied the GLIPH2 algorithm, to cluster TCR sequences into groups that are likely to bind to the same antigen. The GLIPH2 analysis resulted in the identification of 14 Local Similarity Groups, and 26 Global Similarity Groups (Fig.5B). The most significant Local (Motif-Based) and Global (Similarity-based) groups are reported in Table 4 and 5. To infer antigen recognition within the specificity clusters, we assessed their potential recognition of peptides derived from known prostate TAAs: Alpha-methylacyl-CoA racemase (AMACR) [56], PAP[57], PSA [58], PSCA [59], Prostate-specific G-protein coupled receptor (PSGR)[60], PSMA[61], and Protein-glutamine gamma-glutamyltransferase 4 (TGM4) [62]. Peptide recognition was tested using the online tool NetMHCpan with restriction to the most prevalent class I and class II MHC alleles (i.e. HLA-A*01:01; A*02:02; A*03:01; B*07:02; B*15:01; B*08:01; B*15:01; DRB1*01:01; DRB1*01:04) in the Caucasian population[52]. Table 6 summarizes the number of unique TCRβ clonotypes computationally predicted to be associated with each peptide derived from prostate-associated antigens. Notably, PSGR was associated with the highest number of associated TCRβ clonotypes (n=22), followed closely by PSA (n=21) and PSMA (n=17). For each analysed TAA, we selected between 7 and 10 peptides, reaching 67 total peptides for functional validation assays.

**Figure 5:**
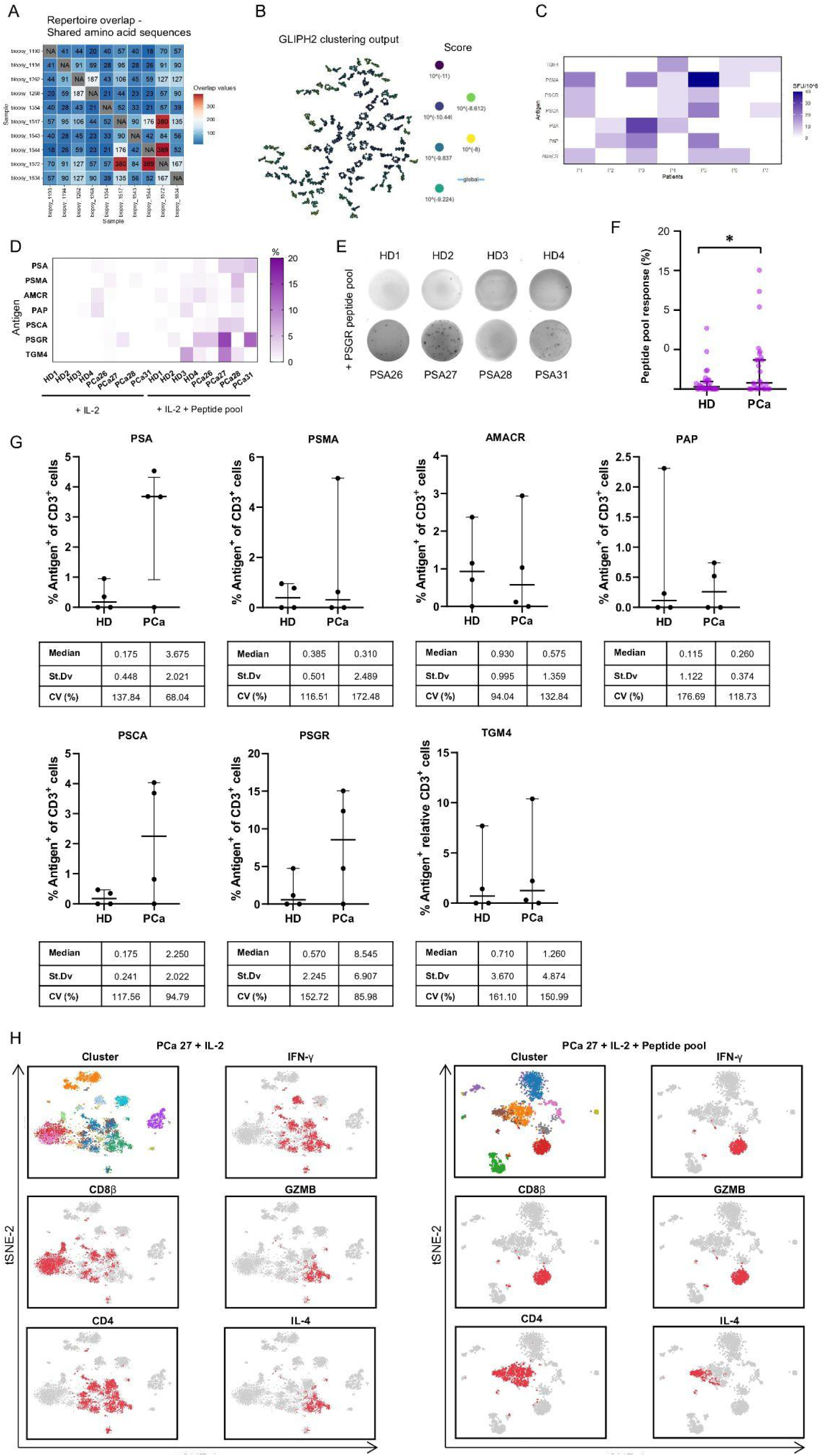
In silico analysis of epitope recognition by shared clonotypes and preliminary functional validation. **A** Matrix showing TILs overlap among patients. **B** GLIPH2 clustering of intratumoral clonotypes. **C** Heatmap representing T-cell response (SFU/10^6) to antigenic pools. **D** Heatmap showing normalized antigen-specific responses for each donor. Values are expressed as the percentage of antigen-induced IFN-γ spots relative to the polyclonal anti-CD3 activator (positive control). **E** Representative IFN-γ ELISpot wells obtained after antigenic expansion with the complete peptide pool and subsequent restimulation with the PSGR-derived peptide pool. Each spot corresponds to a single IFN-γ-secreting cell. **F** Comparison of the overall distribution of normalized peptide-pool responses between healthy donors (HD) and prostate cancer patients (PCa). Each point represents a single donor-peptide combination. Horizontal lines indicate the median and 95% confidence intervals (95% CI). Statistical significance was determined using the Mann–Whitney U test (p = 0.0412). **G** Point plots showing normalized responses to each individual antigenic pool in healthy donors (HD) and prostate cancer patients (PCa). The median standard deviation (SD), and 95% confidence intervals (95% CI) are reported. **H** Single-cell transcriptomic profiling of T-cell populations following expansion in presence of Interleukin-2 (IL-2) only, or IL-2 and peptide pool. t-SNE clustering was used to visualize the expression data. Cells are grouped according to their global gene-expression profiles, and expression patterns of selected genes associated with CD8⁺ and CD4⁺ T-cell states are shown.

**Table 4:**
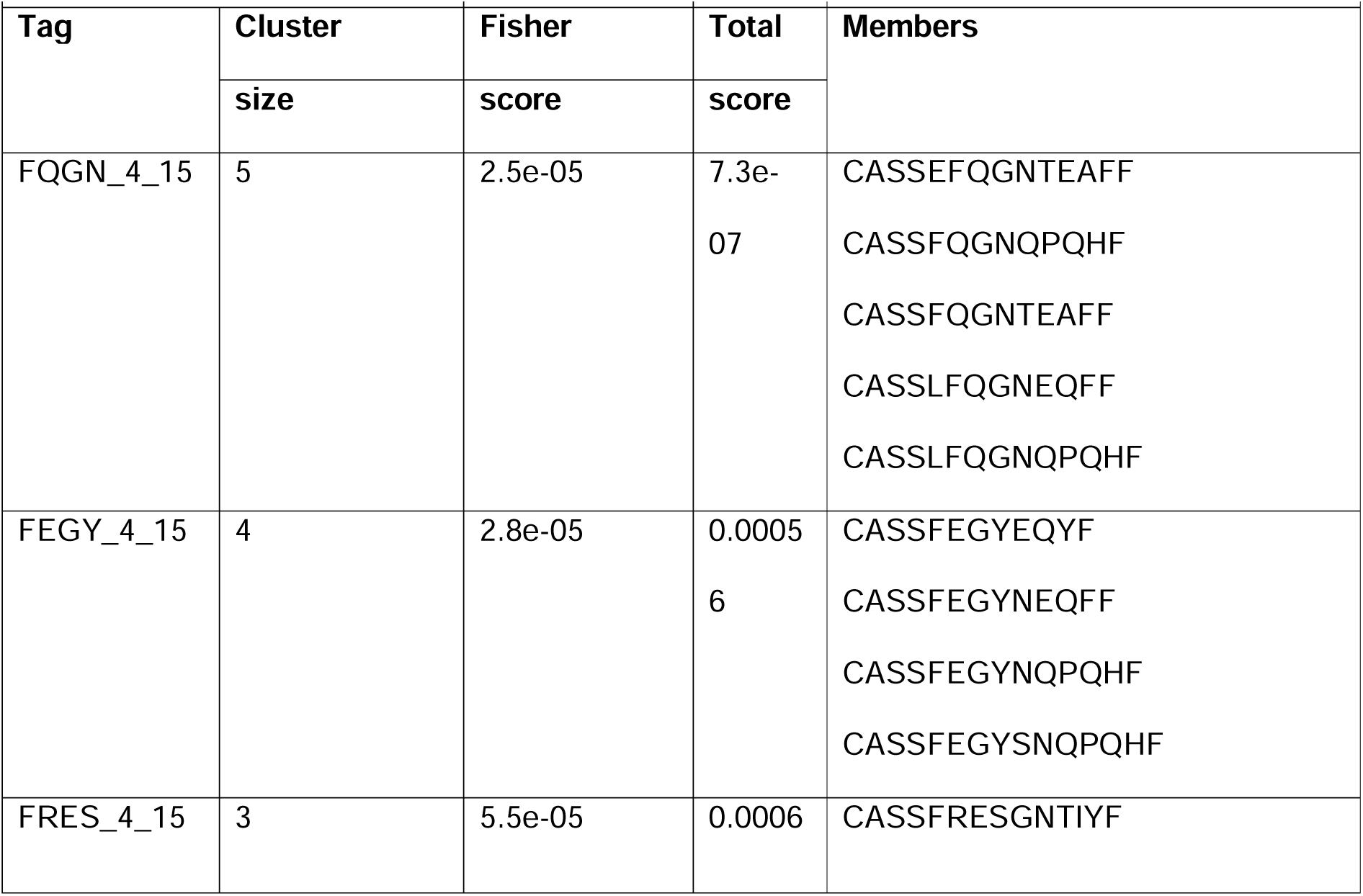
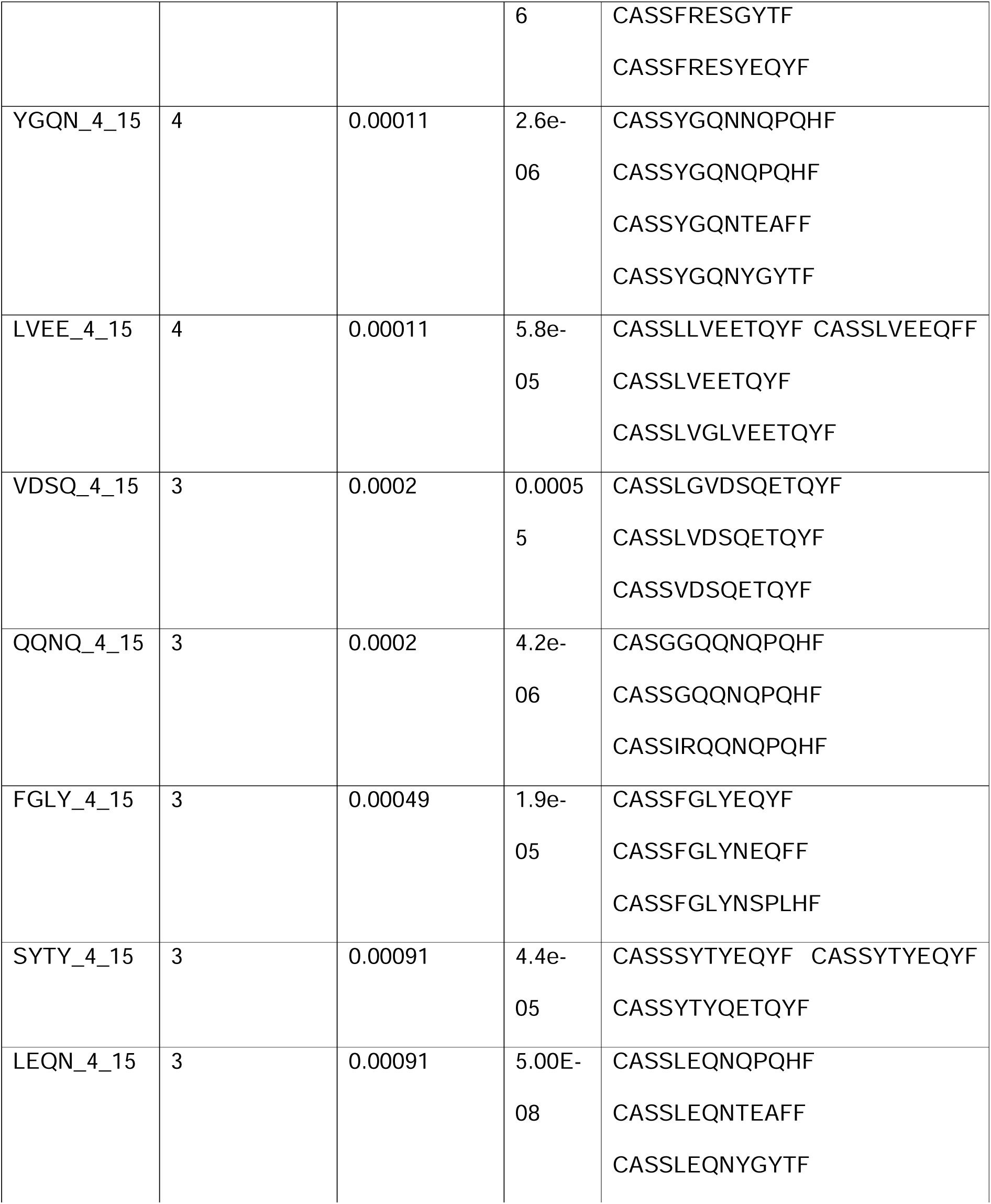
Top Local Specificity Groups.

**Table 5:**
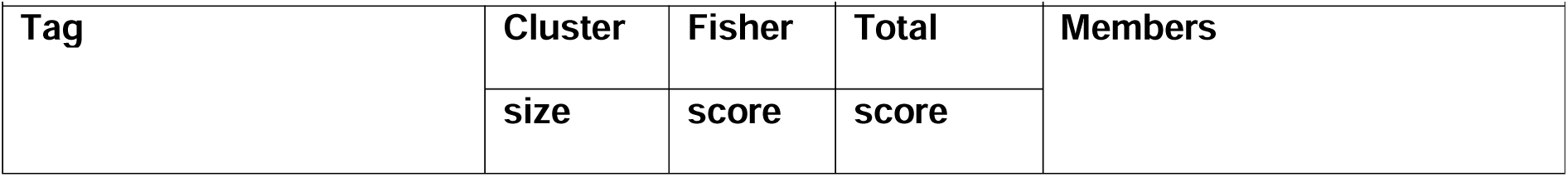
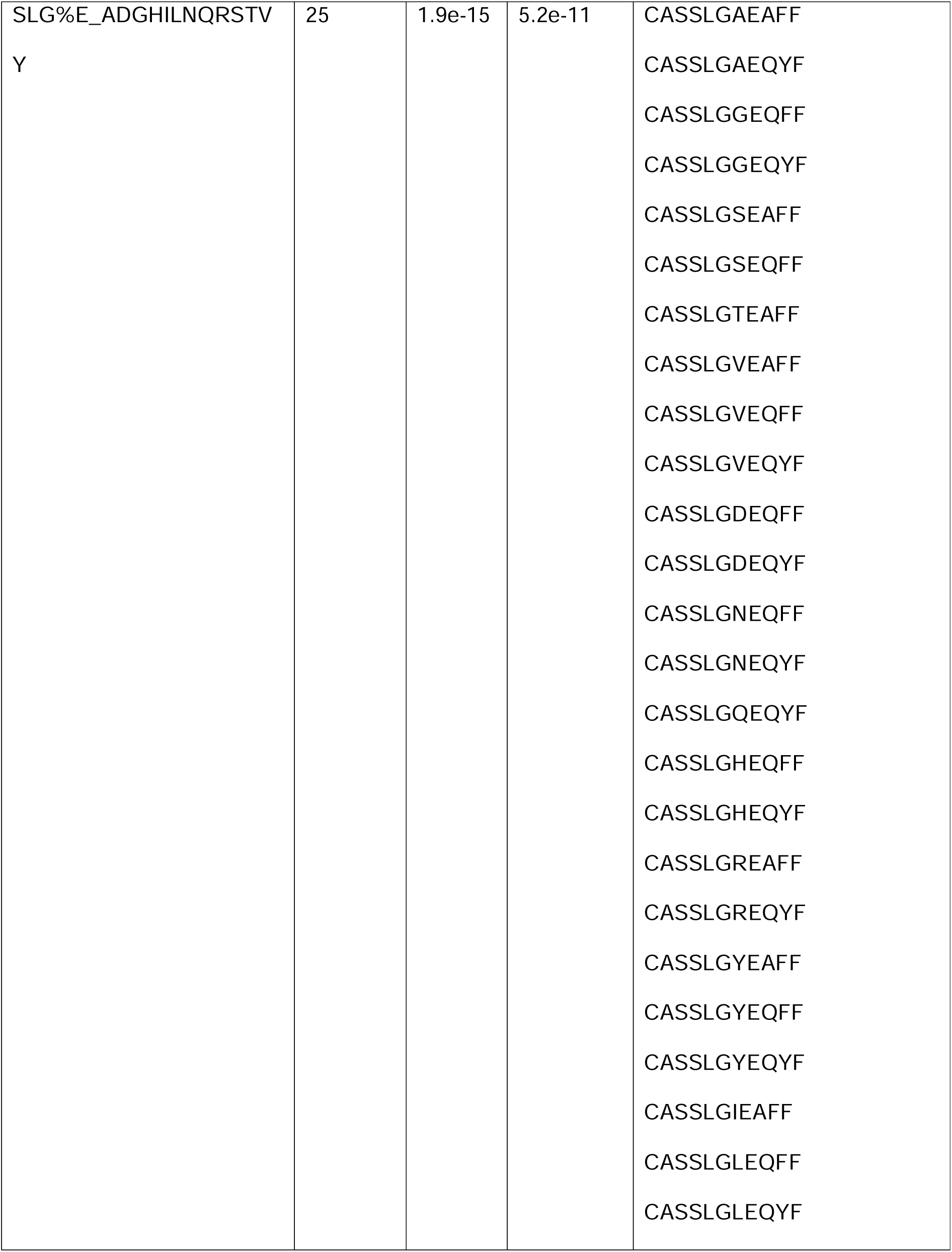
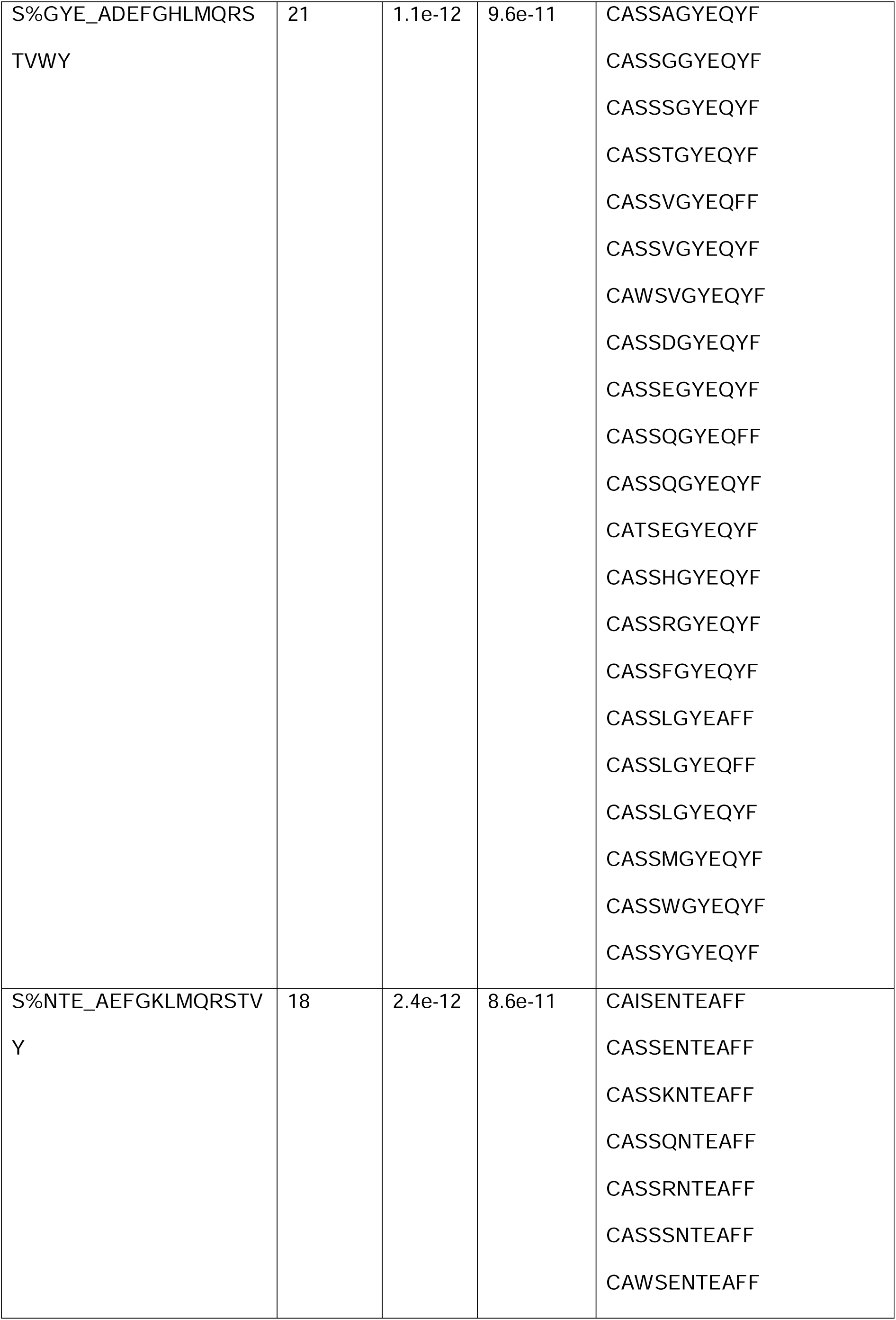
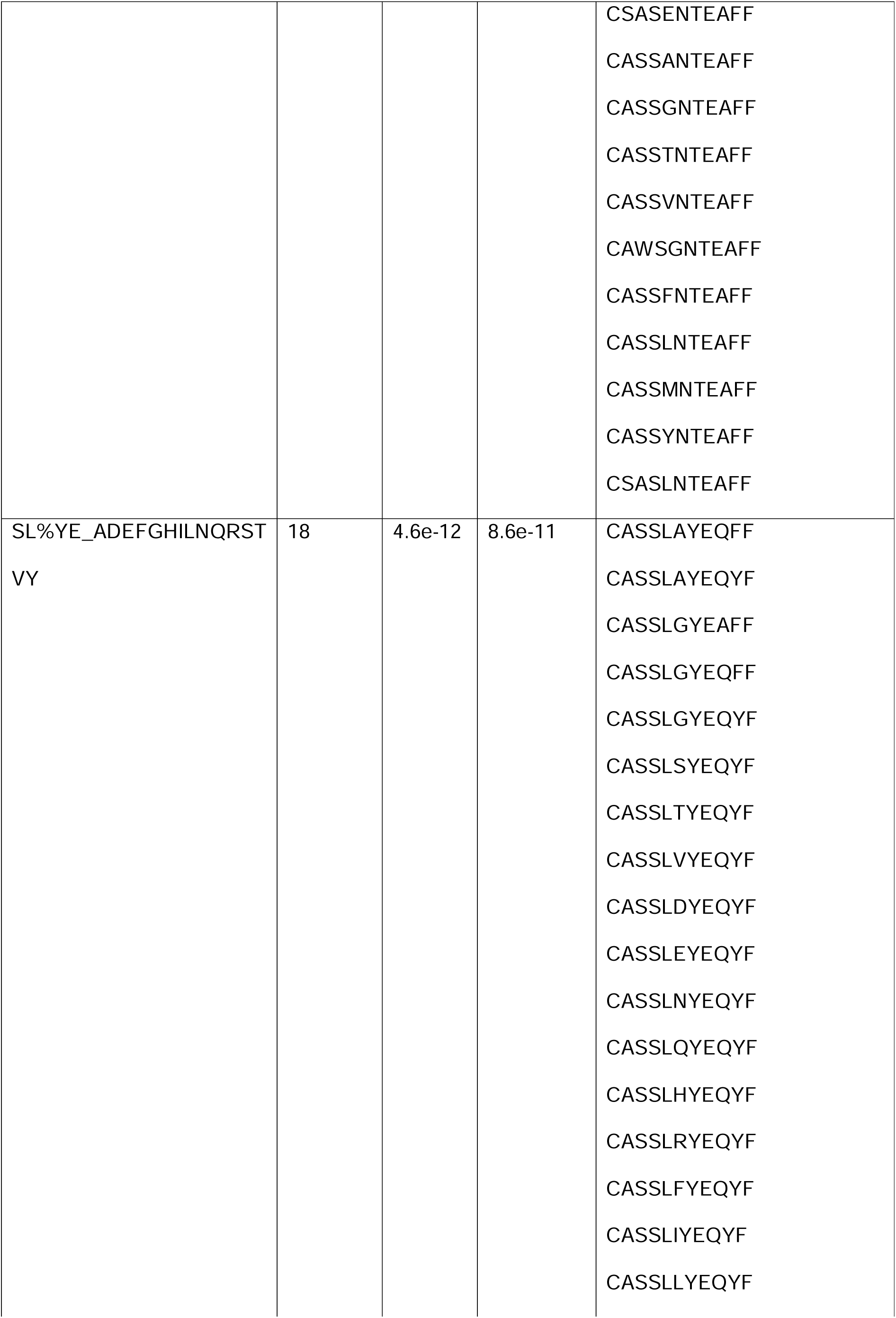
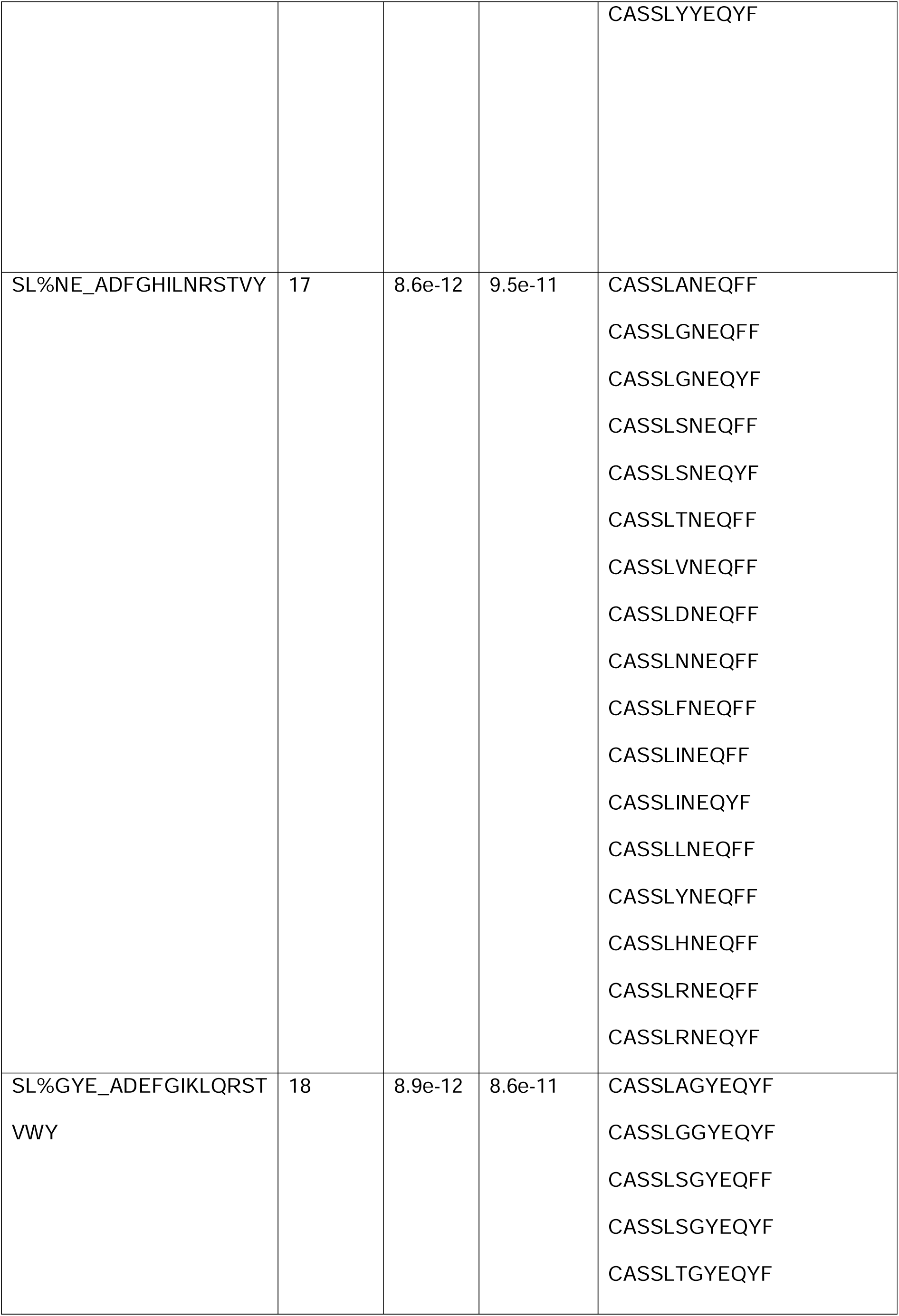
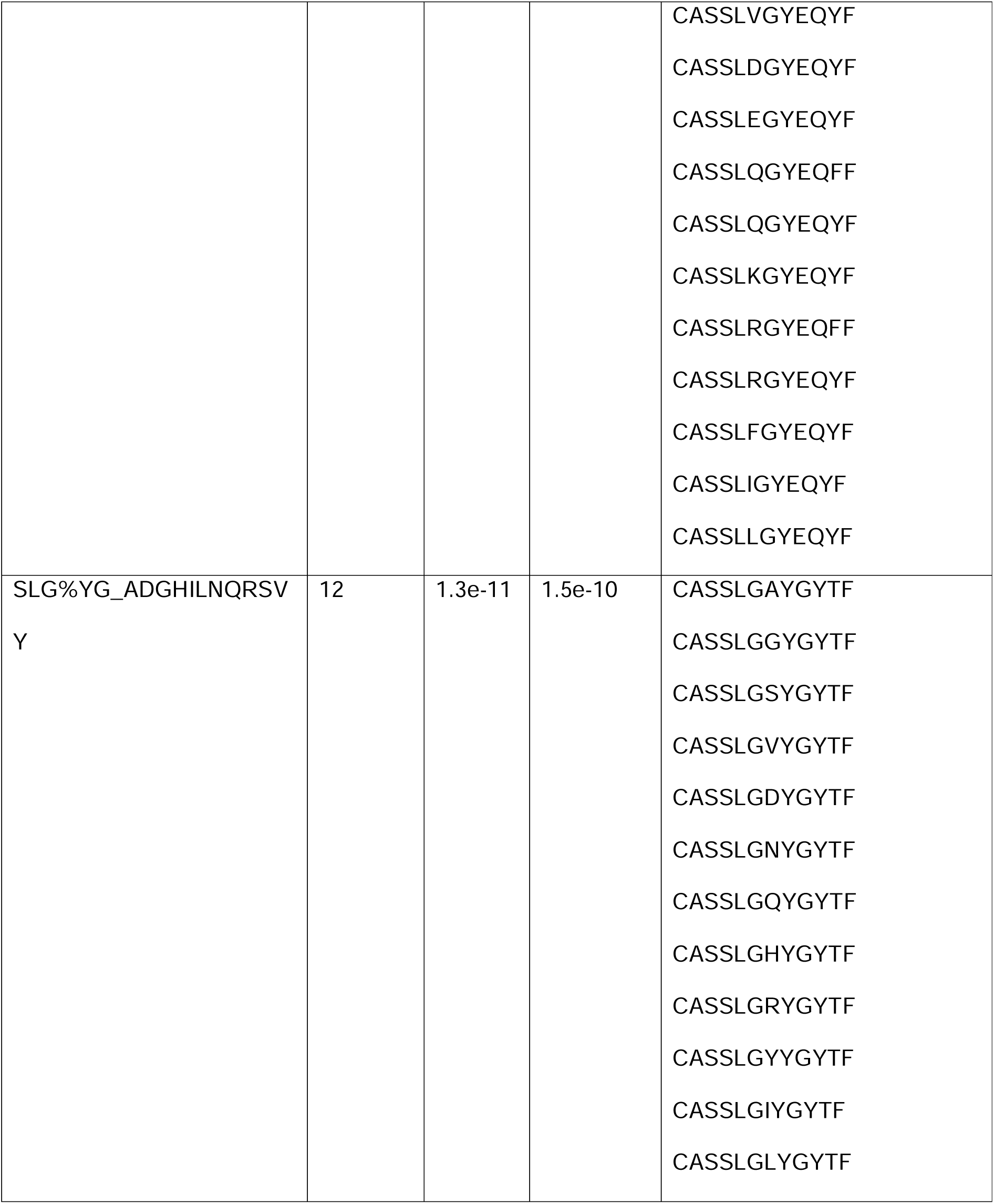
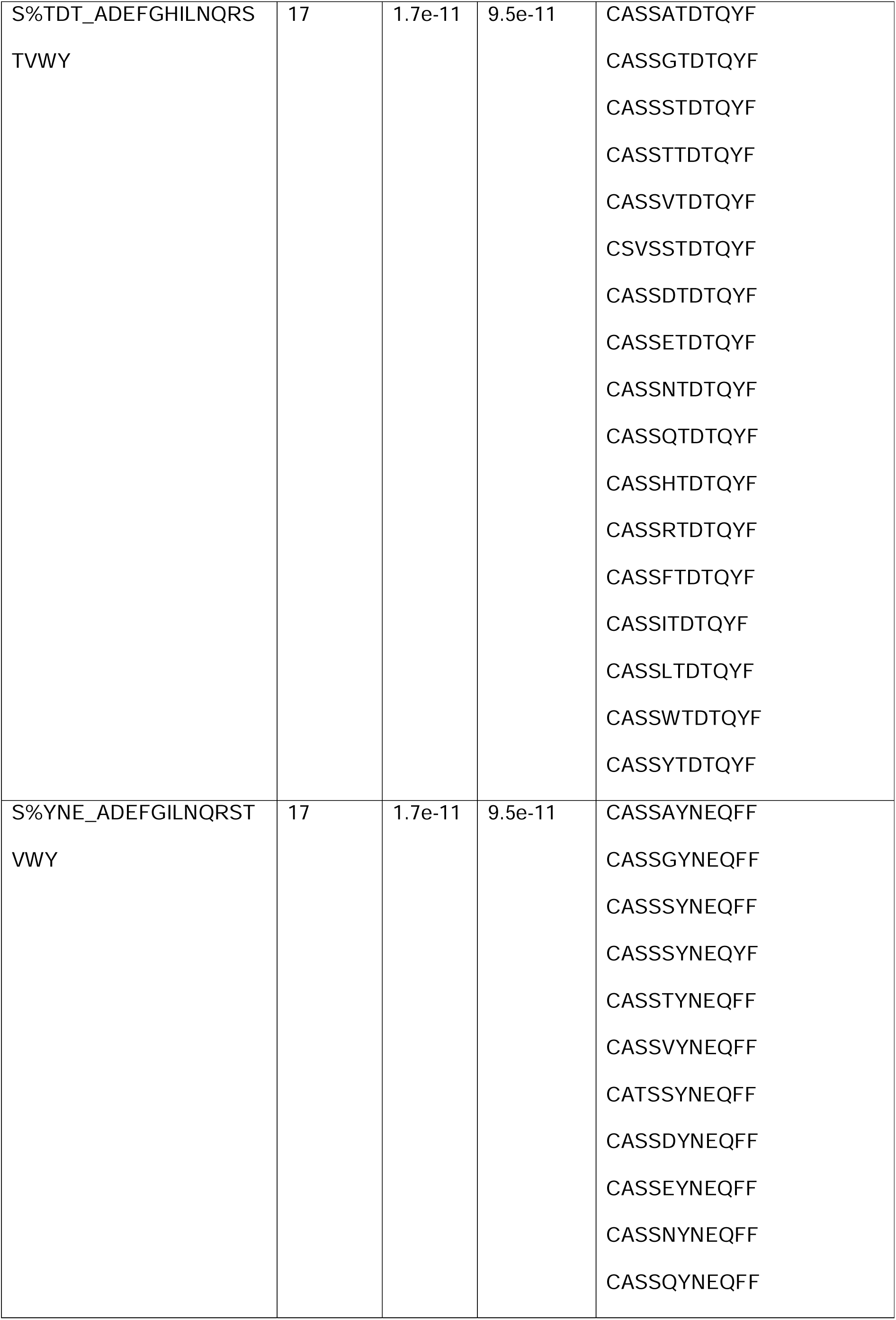
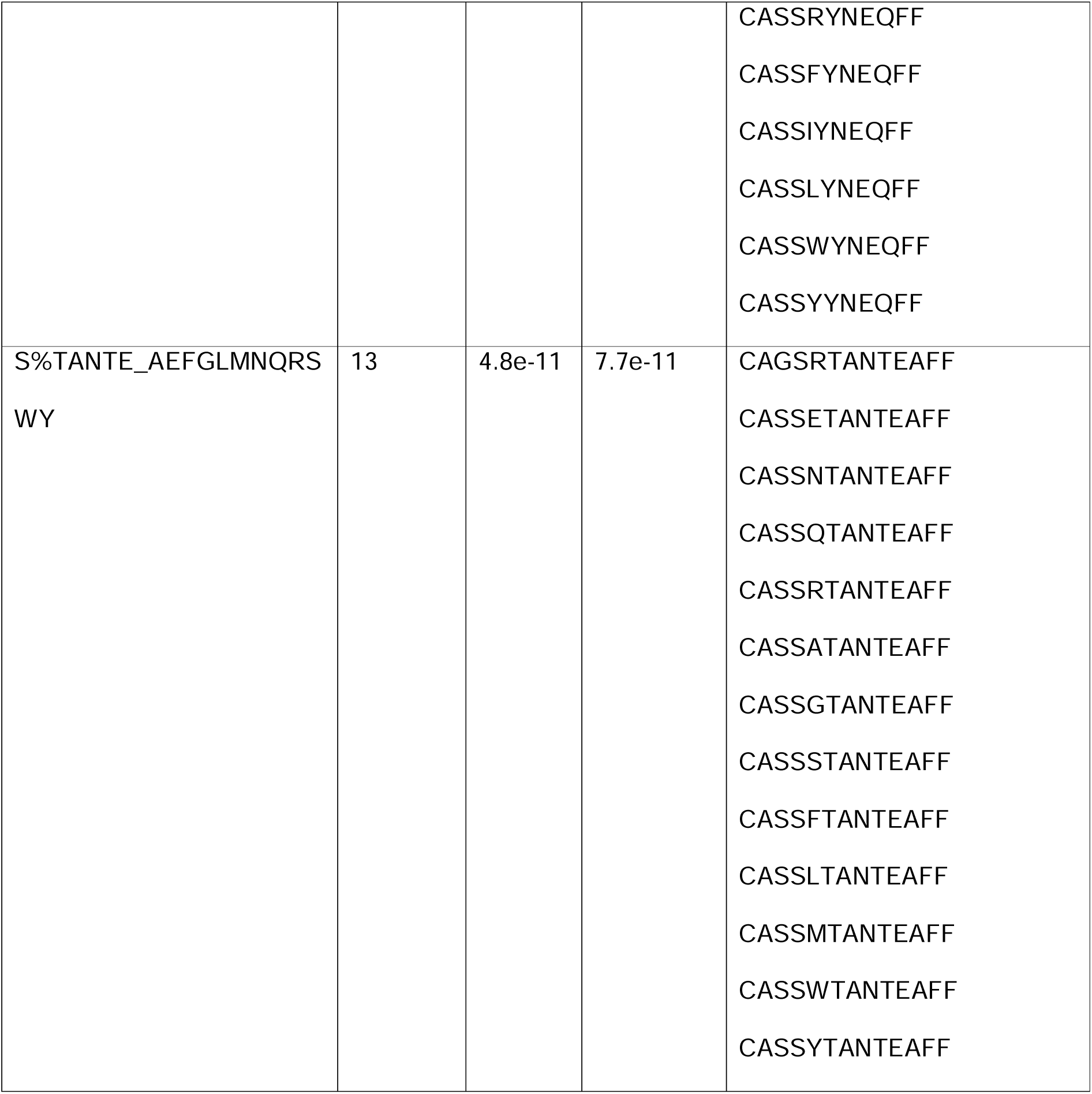
Top Global Specificity Groups.

**Table 6:**
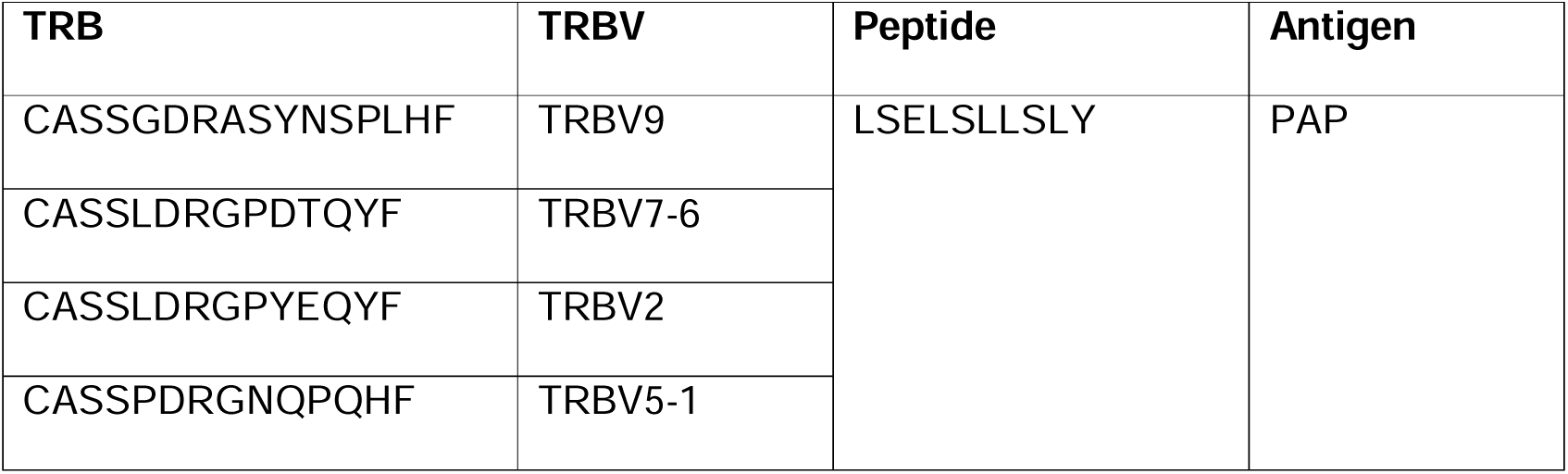
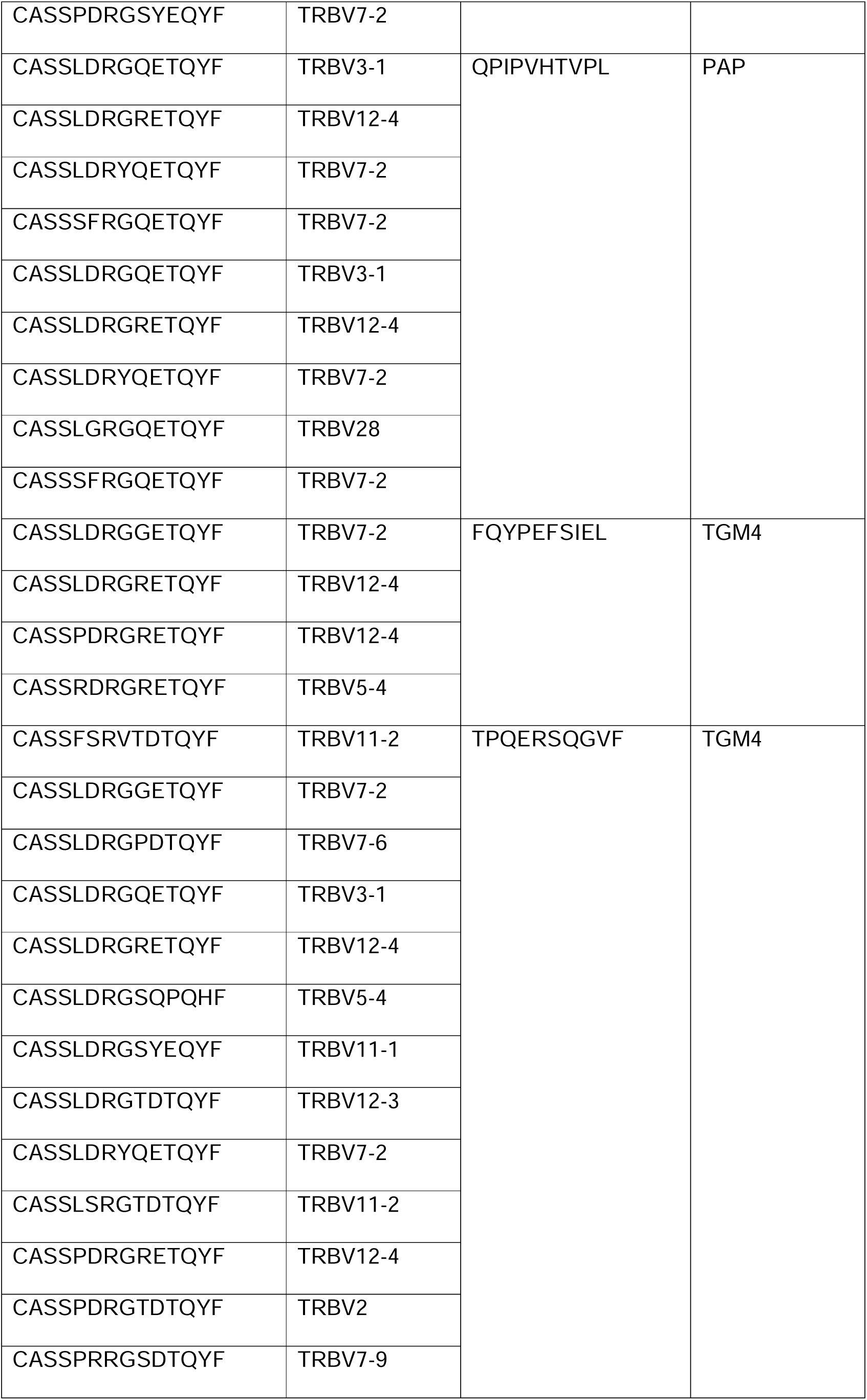
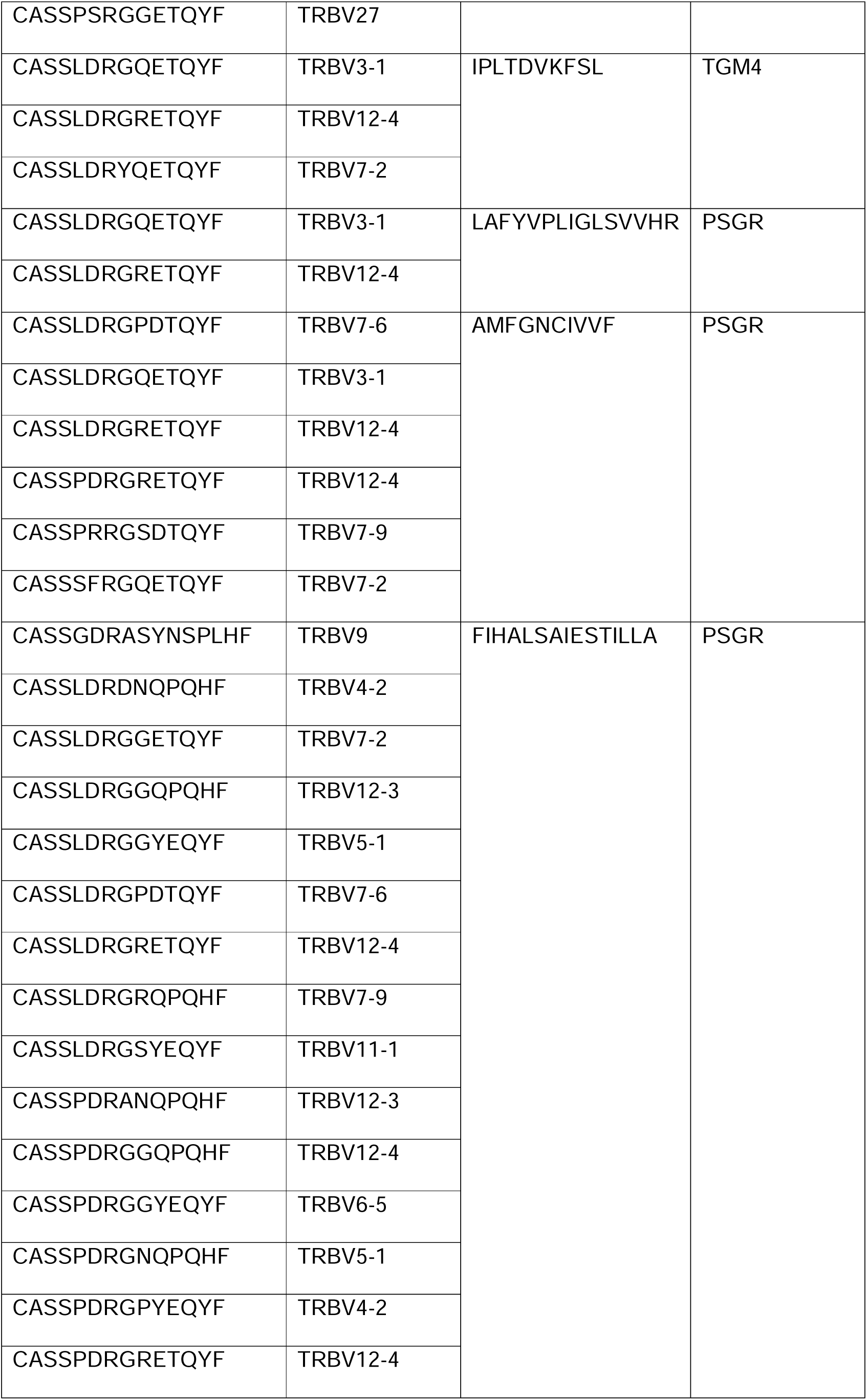
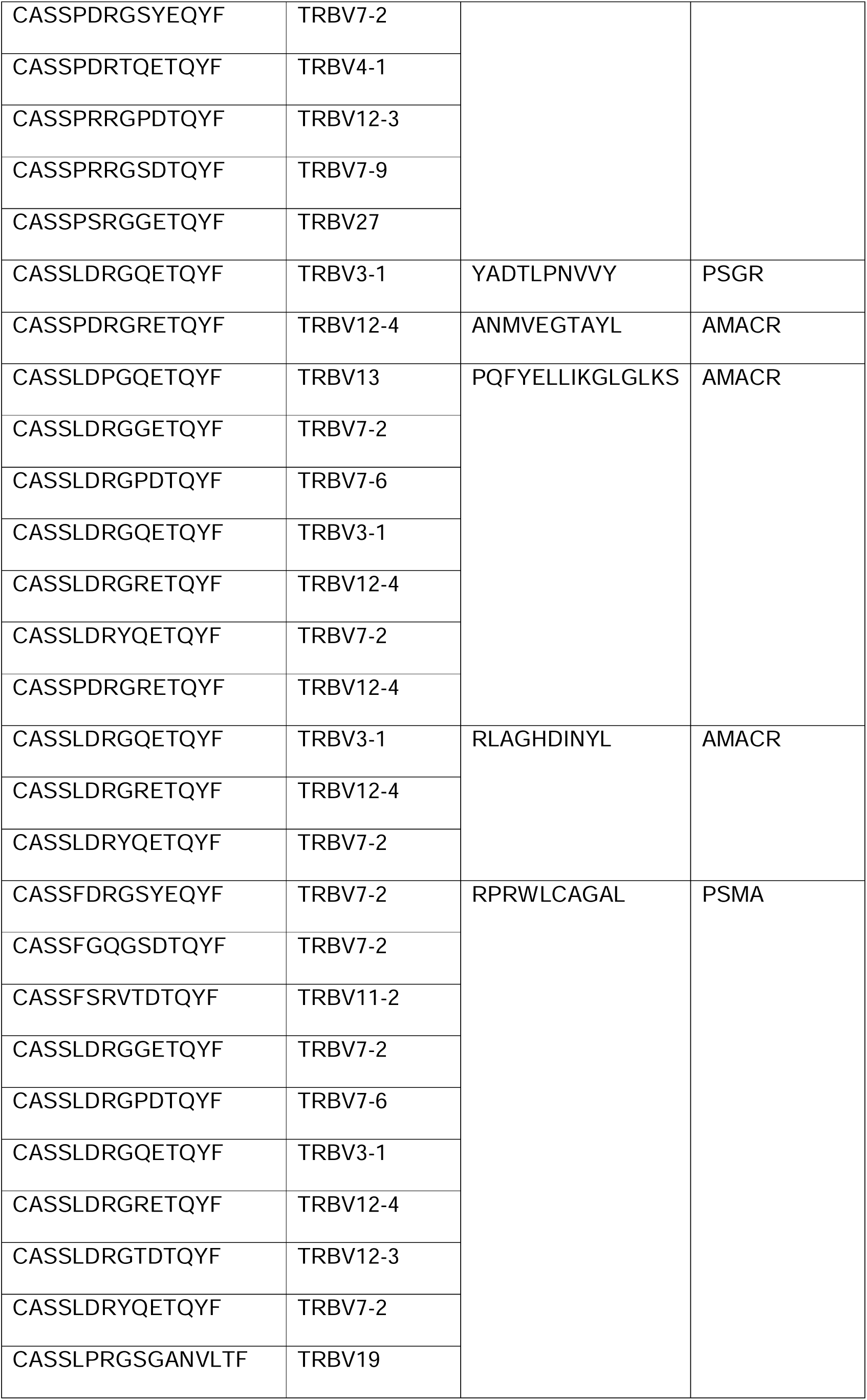
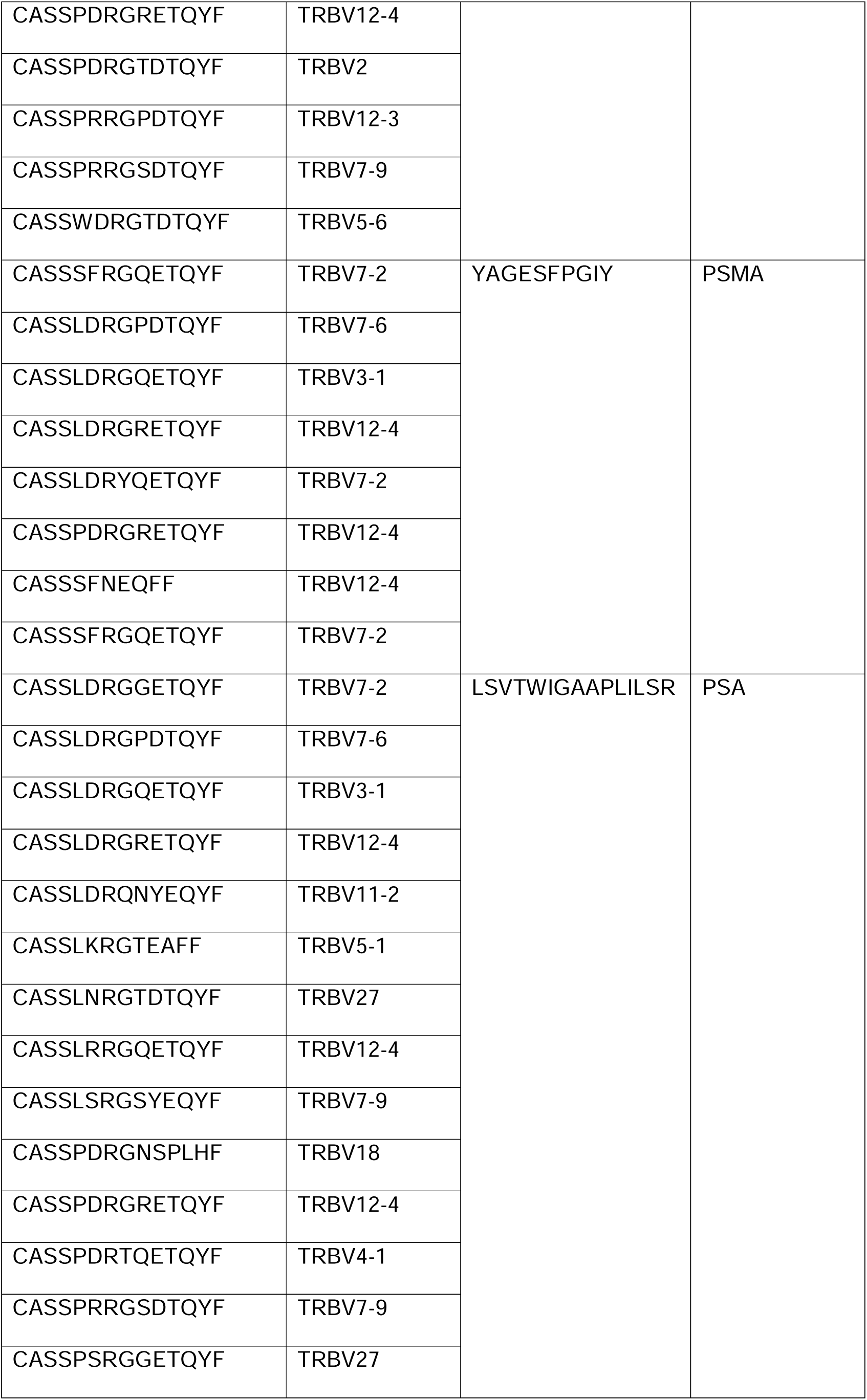
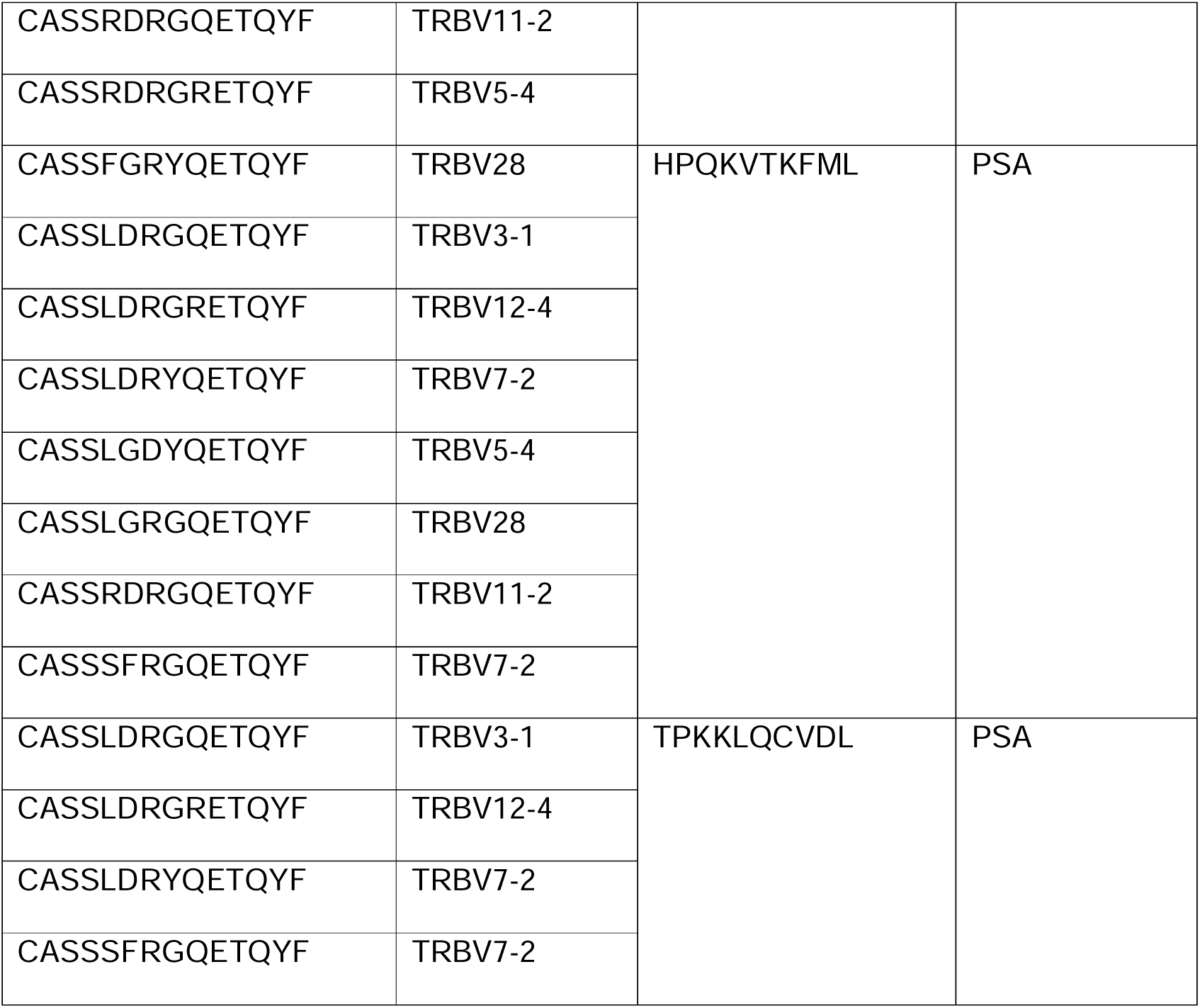
Top T-cell receptor beta (TRB) region, T-cell receptor beta variable region (TRBV) and recognized peptide derived from different Antigens.

### In vitro functional evaluation of candidate peptides

To investigate the biological relevance of the computational predictions, peptide pools derived from the candidate prostate-associated antigens were evaluated using PBMCs from an independent cohort of patients with prostate cancer, owing to the limited availability of material from the discovery cohort.

Ex vivo IFN-γ ELISpot assays showed that all patients responded to at least one antigenic pool (Fig. 5C), although the magnitude of the responses was generally modest (mean 12.60 ± 8.79 SFU/10^6 PBMCs). Among the tested pools, PSMA-derived peptides elicited the highest frequency of responses, being recognized by 71.4% of patients. Because circulating tumor-reactive T cells are expected to be present at low frequency, PBMCs were subsequently expanded for 15 days in the presence of individual peptide pools and IL-2 before reassessing antigen-specific responses by IFN-γ ELISpot (Supplementary Figure 3).

To further evaluate whether these peptide-induced responses were preferentially associated with prostate cancer, peptide-expansion experiments were performed using PBMCs from an independent cohort comprising four PCa patients and four healthy donors. Following expansion with the complete peptide pool, antigen-specific IFN-γ responses were consistently stronger and broader in PCa samples than in healthy donors (Fig. 5D–F). Although no statistically significant differences were observed for individual peptide pools, PSA-, PSCA-, and particularly PSGR-derived peptides showed a trend toward higher responses in PCa patients (Fig. 5G). Given the limited sample size, these findings should be considered supportive rather than definitive evidence of peptide immunogenicity.

To further characterize the cellular populations underlying these responses, two patients from this validation cohort were subsequently analyzed by single-cell RNA/TCR sequencing following expansion in the presence of IL-2 alone or IL-2 plus the peptide pool. The paired TCRαβ sequences corresponding to the ten most abundant clonotypes in the peptide-stimulated condition are reported in Supplementary Table 3. Peptide stimulation was associated with the emergence of a distinct CD8⁺ T-cell cluster expressing IFNG, GZMB, PRF1, CD8A, and CD8B, consistent with an activated cytotoxic transcriptional program (Fig. 5H and Supplementary Fig. 4). A separate activated CD4⁺ population expressing IL2RA, IL4, and IL5 was also identified. These results demonstrate that the candidate peptide pools promote the expansion of discrete activated T-cell populations and provide orthogonal biological support for the computational predictions. However, because these experiments were performed in an independent validation cohort and do not directly establish TCR–peptide–HLA specificity or link the expanded TCRs to those identified in the discovery cohort, the antigen associations described here should be regarded as putative and require further functional validation.

## DISCUSSION

The organization of TCRβ repertoires within tumor-infiltrating lymphocytes (TILs) differs across cancer types, reflecting tumor-specific immune landscapes [24], [63], [64]. Prostate cancer (PCa) represents a paradigmatic immunologically "cold" tumor, characterized by low immunogenicity and an immunosuppressive microenvironment enriched in exhausted T cells, regulatory T cells, myeloid-derived suppressor cells, and M2 macrophages [29], [30] . Although these features limit effective antitumor immunity, previous studies have reported the presence of highly expanded intratumoral TCRβ clonotypes, suggesting that antigen-driven T-cell responses can still emerge despite profound immune suppression [38]. In this context, we characterized matched intratumoral and peripheral blood TCRβ repertoires in ten treatment-naïve PCa patients, revealing distinct clonal architectures, variable overlap, and evidence consistent with repertoire convergence.

We found that intratumoral TCRβ repertoires exhibited reduced richness and increased clonality relative to peripheral blood, extending previous observations of oligoclonal T-cell expansion in prostate cancer tissue [65] . The lower Simpson index and enrichment of medium-to-large clonotype classes in TILs indicate a repertoire architecture characterized by reduced evenness and preferential representation of expanded clonotypes within the tumor compartment. Although this pattern is compatible with local clonal expansion, it should not be interpreted, by itself, as direct evidence of tumor-antigen-driven selection, since peripheral blood contains numerous T-cell populations unrelated to tumor immunity. Moreover, because adjacent benign or histologically normal prostate tissue was not available, repertoire remodeling could only be evaluated relative to peripheral blood rather than to the local non-malignant tissue microenvironment. Similar repertoire constriction has been reported in other solid tumors, including melanoma and renal carcinoma [66], [67], where it has been associated with intratumoral immune responses, including responses directed against tumor-associated antigens and, in some settings, neoantigens. These observations are consistent with previous studies in prostate cancer [38] while emphasizing that the antigenic drivers underlying this repertoire architecture remain to be established.

Although the overall proportion of shared clonotypes between matched TILs and PBMCs did not differ according to Grade Group, complementary frequency-aware overlap metrics suggested differences in repertoire organization. Given the limited cohort size, these findings should be considered exploratory. Using a patient-level summary approach with exact permutation testing, Grade Group ≥2 tumors exhibited a greater absolute number of shared intratumoral clonotypes than Grade Group 1 tumors, whereas normalized overlap metrics showed similar trends but did not consistently reach statistical significance. Comparable patterns of repertoire convergence have been described in melanoma, breast, and lung cancer [68]–[71]. Although our findings do not establish the antigenic basis of this convergence, they are compatible with the hypothesis that common antigenic determinants or shared selective pressures may contribute to shaping the intratumoral T-cell repertoire during prostate cancer progression. Together, these observations highlight the value of integrating complementary overlap metrics, rather than relying on a single index, to characterize repertoire relationships.

To further investigate the antigenic landscape underlying these convergent repertoires, we analyzed the most expanded intratumoral clonotypes using GLIPH2. Multiple specificity groups characterized by conserved CDR3β motifs were identified, several of which contained clonotypes computationally predicted to recognize well-established prostate-associated antigens, including PSMA, PSGR, and PSA. Such motif-level convergence across independent patients is compatible with shared antigen recognition and is consistent with previous large-scale analyses of TCR motif sharing across different cancer types [50], [72]. However, because GLIPH2 and ERGO-II are computational prediction tools and patient-specific HLA typing was not available, these analyses do not establish definitive TCR–peptide–HLA specificity. Instead, they provide a rational framework for prioritizing candidate antigens for functional evaluation.

Consistent with this approach, peptide pools derived from the computationally predicted prostate-associated antigens elicited IFN-γ ELISpot responses and promoted T-cell expansion following in vitro stimulation with IL-2 in an independent validation cohort. Although the ex vivo responses were generally modest, likely reflecting the low precursor frequency of circulating peptide-reactive T cells [73], peptide-expanded cultures exhibited broader and stronger antigen-specific responses in patients with prostate cancer than in healthy donors. Moreover, the identification of clonotypes shared between tumor and peripheral blood suggests that a subset of tumor-associated clonotypes is also detectable in circulation. This finding should not be interpreted as evidence that tumor and peripheral repertoires are equivalent; rather, it supports the concept that circulating T cells may constitute a systemic pool from which selected clonotypes become preferentially enriched within the tumor microenvironment. Preliminary single-cell RNA/TCR sequencing performed on two patients from the validation cohort further identified discrete activated CD8⁺ T-cell populations expressing IFNG, GZMB, and PRF1, together with paired TCRαβ sequences, following peptide stimulation. Although these experiments do not establish direct TCR–peptide–HLA specificity or directly link the expanded TCRs to the clonotypes identified in the discovery cohort, they provide orthogonal evidence that the candidate peptide pools promote the expansion of activated cytotoxic T cells. Among the evaluated antigens, PSGR was associated with the largest number of putative TCRβ clonotypes and consistently supported in vitro T-cell expansion, suggesting that it represents a particularly promising candidate for future T-cell-based immunotherapies. Despite compelling preclinical evidence supporting its immunogenicity [74], no clinical studies specifically targeting PSGR have yet been reported.

Importantly, the presence of expanded intratumoral clonotypes, including those computationally predicted to recognize prostate-associated antigens, should not be interpreted as evidence of effective immune surveillance. Rather, our findings reinforce the concept that PCa retains the capacity to prime antigen-specific T-cell responses while simultaneously maintaining a profoundly immunosuppressive tumor microenvironment that limits their effector function. This immune dysfunction is likely mediated by multiple complementary mechanisms, including T-cell exhaustion, impaired antigen presentation, insufficient costimulatory signaling, and the accumulation of regulatory T cells, M2 macrophages, and myeloid-derived suppressor cells [29], [30]. Consequently, local clonal expansion and candidate antigen-specific immune responses can coexist with ineffective tumor control, a defining feature of immunologically cold tumors.

Several limitations should be acknowledged. First, the study included only ten treatment-naïve PCa patients, limiting subgroup analyses and increasing susceptibility to selection bias, although the cohort was sufficient to identify recurrent repertoire features and generate biologically plausible hypotheses. Second, adjacent benign tissue and immunohistochemical characterization were not available. Consequently, repertoire remodeling could only be assessed relative to peripheral blood, and the density, spatial distribution, and phenotype of infiltrating immune cells could not be evaluated. Third, antigen specificity was inferred using computational approaches such as GLIPH2 and ERGO-II, which identify putative shared specificity groups based on TCR sequence similarity rather than direct TCR–peptide–HLA interactions. In addition, patient-specific HLA typing was not available for either the discovery or the validation cohort. Therefore, these analyses provide hypothesis-generating evidence rather than definitive specificity assignments. Fourth, the functional peptide-stimulation experiments and single-cell RNA/TCR sequencing were performed in an independent validation cohort. Although these analyses provide complementary evidence supporting the biological relevance of the candidate peptides and demonstrate the expansion of activated T-cell populations following peptide stimulation, they cannot directly establish the antigen specificity of the clonotypes identified in the discovery cohort or demonstrate definitive TCR–peptide–HLA recognition. Moreover, paired αβ TCR sequencing was available for only two patients, limiting the generalizability of these observations.

Future studies should integrate patient-specific HLA typing, paired single-cell TCRαβ sequencing, spatial characterization of the tumor microenvironment, and direct functional validation to define antigen specificity and evaluate the therapeutic potential of candidate clonotypes.

## CONCLUSIONS

Treatment-naïve prostate cancer exhibits restricted intratumoral TCRβ repertoires enriched for expanded clonotypes that partially overlap with the peripheral blood repertoire. Computational prediction, functional peptide-stimulation assays, and preliminary single-cell RNA/TCR sequencing collectively support the biological relevance of candidate prostate-associated antigens, although definitive TCR–peptide–HLA specificity remains to be established. These findings provide a framework for the future development of TCR-based biomarkers and antigen-directed immunotherapies in prostate cancer.

## DATA AVAILABILITY

Accession number SRX33156384: https://www.ncbi.nlm.nih.gov/sra/SRX33156384[accn];

Accession number SRX33156385: https://www.ncbi.nlm.nih.gov/sra/SRX33156385[accn];

Accession number SRX33156386: https://www.ncbi.nlm.nih.gov/sra/SRX33156386[accn];

Accession number SRX33156387: https://www.ncbi.nlm.nih.gov/sra/SRX33156387[accn];

Accession number SRX33156388: https://www.ncbi.nlm.nih.gov/sra/SRX33156388[accn];

Accession number SRX33156389: https://www.ncbi.nlm.nih.gov/sra/SRX33156389[accn];

Accession number SRX33156390: https://www.ncbi.nlm.nih.gov/sra/SRX33156390[accn];

Accession number SRX33156391: https://www.ncbi.nlm.nih.gov/sra/SRX33156391[accn];

Accession number SRX33156392: https://www.ncbi.nlm.nih.gov/sra/SRX33156392[accn];

Accession number SRX33156393: https://www.ncbi.nlm.nih.gov/sra/SRX33156393[accn];

Accession number SRX33156394: https://www.ncbi.nlm.nih.gov/sra/SRX33156394[accn];

Accession number SRX33156395: https://www.ncbi.nlm.nih.gov/sra/SRX33156395[accn];

Accession number SRX33156396: https://www.ncbi.nlm.nih.gov/sra/SRX33156396[accn];

Accession number SRX33156397: https://www.ncbi.nlm.nih.gov/sra/SRX33156397[accn];

Accession number SRX33156398: https://www.ncbi.nlm.nih.gov/sra/SRX33156398[accn];

Accession number SRX33156399: https://www.ncbi.nlm.nih.gov/sra/SRX33156399[accn];

Accession number SRX33156400: https://www.ncbi.nlm.nih.gov/sra/SRX33156400[accn];

Accession number SRX33156401: https://www.ncbi.nlm.nih.gov/sra/SRX33156401[accn];

Accession number SRX33156402: https://www.ncbi.nlm.nih.gov/sra/SRX33156402[accn];

Accession number SRX33156403: https://www.ncbi.nlm.nih.gov/sra/SRX33156403[accn].

## ABBREVATIONS

TCRß: T-cell receptor beta chain
CDR3: Complementarity-determining region 3
PCa: Prostate Cancer
PBMCs: Peripheral Blood Mononuclear Cells
TILs: Tumor-infiltrating Lymphocytes
TAAs: Tumor associated antigens
PSMA: Prostate Specific Membrane Antigen
PSA: Prostate Specific Antigen
PSGR: Prostate-specific G-protein coupled receptor
AMACR: Alpha-methylacyl-CoA racemase
TGM4: Protein-glutamine gamma-glutamyltransferase 4

## ACKNOWLEDGEMENTS

I.A. is supported by the Next Generation EU - Italian NRRP, Mission 4, Component 2, Investment 1.5, call for the creation and strengthening of ’Innovation Ecosystems’, building ’Territorial R&D Leaders’ (Directorial Decree n. 2021/3277) - project Tech4You - Technologies for climate change adaptation and quality of life improvement, n. ECS0000009.

R.G. is supported by Fondazione Umberto Veronesi and Fondazione Ferrero.

## Contributions

A.Ab., C.G. Methodology, Investigation. I.R.L., M.C.S., Methodology. G.F., D.M., R.D., F.Ca., G.C., Methodology, Supervision. S.A., A.Am., Clinical Sample Collection. A.P., C.F., and F.Cr. Data curation. R.G. and I.A. Methodology, Investigation, Data analysis, Writing-Original draft preparation. C.P. Funding acquisition, Conceptualization, Supervision, Writing-review and editing.

Antonio Abatino, Caterina Giordano, and Lisa Isdraele Romano contributed equally.

Ilenia Aversa and Camillo Palmieri contributed equally.

## DECLARATIONS

### Ethical approval

This study is a retrospective study and was approved by the Regional Ethical Review board of Regione Calabria (Ethics committee approval number: 84/2026; Clinical trial number: not applicable). The study was conducted in accordance with the Italian National regulations (Article 110 of Legislative Decree 196/2003 and its amendments, including Legislative Decree 19/2024), the Declaration of Helsinki, and EU Regulation 2016/679 (GDPR). The study used irreversibly anonymized surplus diagnostic material that could not be linked to identifiable individuals.

### Consent to participate

All participants provided written informed consent authorizing the use of their anonymized biological material and associated clinical data for research purposes, at the time of sample collection.

### Consent to publish

Not applicable.

